# Integrated mapping of human meniscus and cartilage eQTLs reveals shared and distinct osteoarthritis genetic drivers

**DOI:** 10.64898/2026.04.12.26350702

**Authors:** Yutaro Uchida, Yuta Fujii, Hannah Swahn, Mahoko Takahashi Ueda, Tomoki Chiba, Takahide Matsushima, Yuki Naito, Ryo Nakamichi, Kenji Takahashi, Merissa Olmer, The RE-JOIN Consortium Investigators, Martin Lotz, Yuta Kochi, Hiroshi Asahara

## Abstract

Osteoarthritis (OA) is a prevalent musculoskeletal disorder and a leading cause of global disability. Although meniscal damage is a major risk factor of OA pathogenesis, genetic regulatory studies have remained largely confined to articular cartilage. Here, we establish the first comprehensive expression quantitative trait locus (eQTL) map integrating whole-genome sequencing and bulk transcriptomics from human meniscus (n=112) and cartilage (n=113). Supported by single-nucleus multiomics (cartilage: 56,549 nuclei; meniscus: 34,343 nuclei), we uncovered highly tissue-specific genetic risk architectures. Colocalization with OA GWAS identified 27 meniscus-specific, 28 shared, and 20 cartilage-specific causal genes. Chromatin-informed fine-mapping and deconvolution elucidated distinct pathogenic mechanisms; notably, meniscus-specific signals converged on *VEGFA* via rare promoter variants and an enhancer in fibrochondrocyte progenitors, alongside a shared eQTL for *CLEC18A*. Exploratory analysis suggested candidate compounds to reverse pathogenic gene expression. Our findings underscore the meniscus as a distinct genetic driver, molecularly reinforcing OA as an entire joint organ failure.

## Introduction

Osteoarthritis (OA) is one of the most prevalent musculoskeletal disorders in our aging society, posing a critical public health challenge that affects approximately 40% of individuals over the age of 65 and more than 200 million people worldwide^1–3^. Although OA has long been described primarily as “wear and tear” or degenerative changes of the articular cartilage, the concept has evolved significantly in recent years. It is now viewed as a “whole-joint disease” involving complex interactions among multiple joint tissues, including cartilage, meniscus, subchondral bone, synovium, infrapatellar fat pad, tendons, and ligaments^4,5^.

Parallel to this conceptual evolution, recent large-scale genome-wide association studies (GWAS) have substantially advanced our understanding of the genetic basis of OA. Multiple studies, including meta-analyses involving over 2 million individuals, have identified more than 1,500 loci associated with OA risk^6,7^, leaving no doubt that OA has a strong genetic component. However, the majority of these disease-associated variants are located in non-coding regions, and it remains largely unclear through which tissues, cell types, or molecular mechanisms they contribute to disease risk^6,7^.

Expression quantitative trait locus (eQTL) analysis is a powerful method for dissecting the effects of GWAS-identified single nucleotide variants (SNVs) on gene expression in specific tissues^8–11^. In the context of OA, eQTL analyses have predominantly focused on cartilage.

Indeed, studies utilizing low-grade and high-grade OA cartilage, as well as multi-omics approaches combining methylation profiles, chromatin structure, ATAC-seq, and Hi-C, have identified multiple eQTL signals colocalizing with OA GWAS loci^9–11^. Furthermore, analyses using primary chondrocytes under inflammatory stimulation have reported alterations in gene regulation that mimic disease conditions^8^.

The meniscus plays a role as critical as cartilage in OA pathology. Composed of an avascular, cartilage-like inner zone and a vascular, fibrocartilaginous outer zone, the meniscus functions in load distribution, shock absorption, and joint stabilization. Epidemiologically, meniscal injury and meniscectomy are established strong risk factors for OA onset. Pathologically, the meniscus undergoes calcification, fibrosis, hypertrophy-like changes, and activation of inflammatory signals during OA progression^12^. Nevertheless, systematic analyses of how genetic variants in the meniscus affect gene regulation and contribute to OA risk have been lacking.

While damage to meniscus and cartilage is closely correlated and a main driver of OA initiation and progression, eQTL analyses targeting the meniscus and their integration with OA GWAS data have not yet been reported. Furthermore, although previous eQTL studies have investigated OA cartilage^8,10^, their reliance on diseased tissues limits the evaluation of genetic variants in normal states. Consequently, it remains unknown how OA-associated variants influence gene regulation in the normal state and how these effects are altered with disease initiation and progression. In addition, no study has simultaneously analyzed chromatin state and gene expression at the single-nucleus level in any of the joint tissues, leaving the impact of cell-type-specific chromatin structural differences on eQTL effects unknown.

Against this background, we conducted a tissue-specific and cell-type-specific eQTL analysis of both cartilage and meniscus, including normal specimens, by combining whole-genome sequencing (WGS) and bulk transcriptome analysis with single-nucleus multiomics. Furthermore, by integrating deconvolution analysis, enhancer prediction, and fine-mapping, we aimed to identify regulatory variants colocalizing with OA GWAS signals based on multi-omic evidence. We also utilized Promoter AI^13^ to investigate the impact of rare variants on gene expression. Finally, leveraging these genetically anchored transcriptomic signatures, we performed drug response prediction to identify candidate compounds capable of reversing the pathogenic networks in both cartilage and meniscus. This study expands the genetic basis of OA from a cartilage-centric view towards the whole joint, and by elucidating gene regulatory abnormalities at the multi-tissue and multi-cell-type level presents a new understanding and potential classification of OA pathology.

## Results

### Overview of multi-omics analysis using cartilage and meniscus specimens

To conduct eQTL analysis in meniscus and cartilage, we collected matched cartilage and meniscus samples from the same tissue donors with normal (Outerbridge Grade 0-1; cartilage n=41, meniscus n=43), mild OA (Grade 1.5-3; cartilage n=18, meniscus n=18), or from patients undergoing total knee arthroplasty (TKA) for severe OA (Grade4; cartilage n=54, meniscus n=51) and performed bulk RNA-seq and whole-genome sequencing (WGS) on all specimens. Based on the principal component analysis (PCA), the samples were predicted to consist of 87 European (EUR), 24 American (AMR), and 7 African (AFR) ancestry individuals (**Supplementary Fig. S1, Supplementary Table S1**). Furthermore, to enable cell-type-specific analyses, we performed single-nucleus multiomics analysis on a subset of cartilage (normal: n=12, severe OA n=6) and meniscus (normal: n=6, severe OA: n=7) tissues. Using these data, we estimated cell-type-specific enhancer regions using SCENT^14^ and deconvolved bulk RNA-seq data using Bayes Prism^15^ to estimate gene expression levels for each cell type. Based on these integrated datasets, we conducted eQTL analyses for both bulk and cell-type-specific gene expression, followed by colocalization analysis with OA GWAS SNVs and fine-mapping. Integrating these multi-layered genomic insights, we identified potential therapeutic compounds capable of reversing GWAS-linked eQTL effects and transcriptomic signatures associated with OA pathology (**Fig. 1a**).

**Fig. 1.**
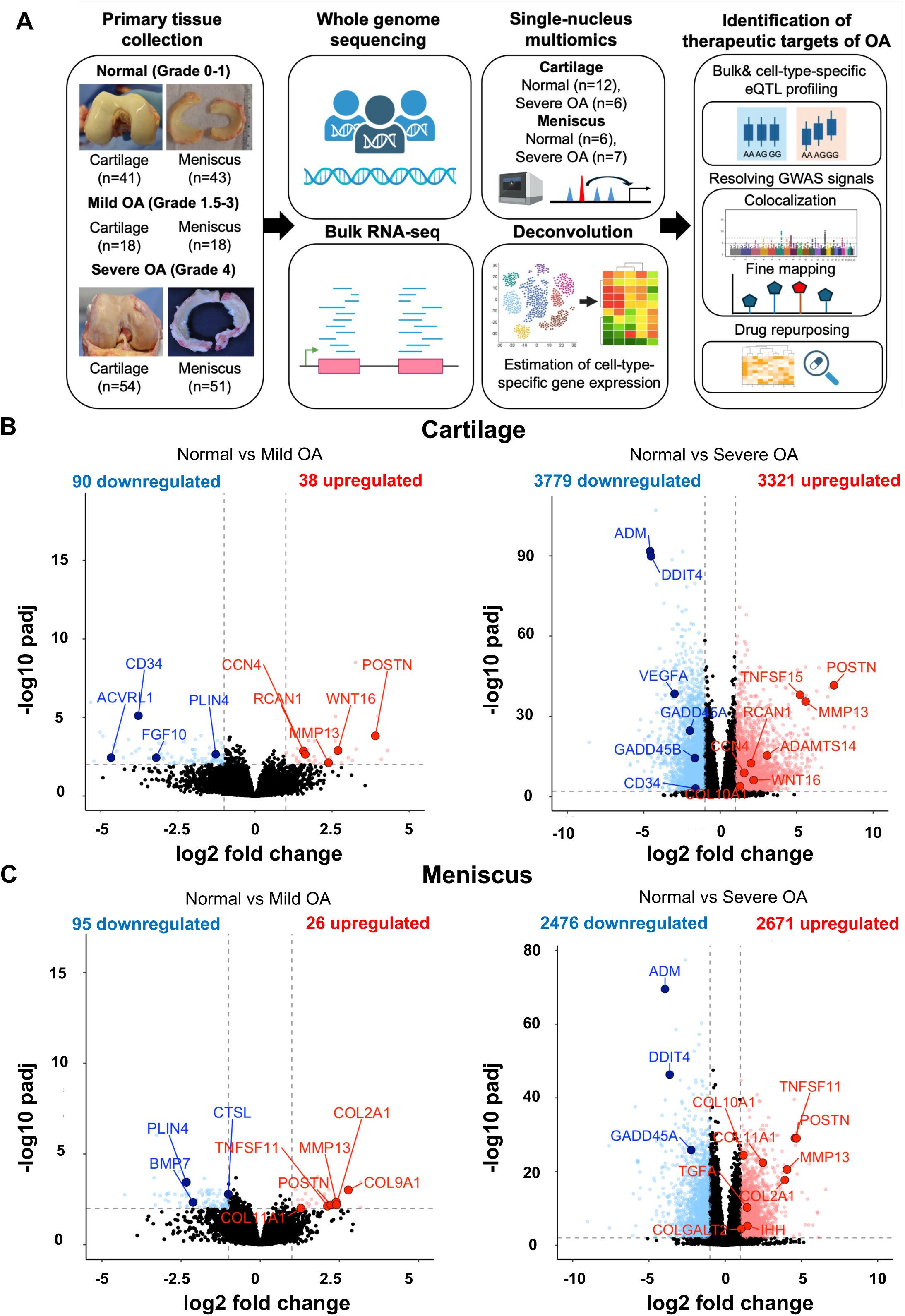
Integrated multiomic profiling of cartilage and meniscus in osteoarthritis. A, Overview of the study design. Human articular cartilage and meniscus samples spanning normal, mild osteoarthritis (OA), and severe OA states were subjected to bulk RNA sequencing and whole-genome sequencing. A subset of samples was further profiled using single-nuclei multiome sequencing to jointly capture chromatin accessibility and gene expression. Bulk transcriptomic data were deconvolved using single-cell references, and integrated analyses included bulk and cell-type–specific eQTL mapping, colocalization with OA GWAS signals, and chromatin-informed fine-mapping. **B,** Principal transcriptional changes associated with OA in cartilage (left) and meniscus (right). Volcano plots show differentially expressed genes (DEGs) between normal (grade 0–1) and severe OA (grade 4) samples. Red dots indicate significantly upregulated genes (log2 fold change > 1, adjusted *P* < 0.05), and blue dots indicate downregulated genes (log2 fold change <-1, adjusted *P* < 0.05). Representative OA-related genes are labeled.

### Transcriptome analysis in cartilage and meniscus

Comparative analysis with mild OA tissues highlighted critical early-stage pathological markers. We found that *MMP13* and *POSTN* were commonly upregulated in both cartilage and meniscus. Notably, the meniscus exhibited a distinct signature characterized by the specific upregulation of collagen genes, such as *COL2A1* and *TNFSF11* (*RANKL*). As the disease progressed to severe OA, bulk RNA-seq revealed more extensive transcriptomic changes, with 3,321 upregulated and 3,779 downregulated genes in cartilage and 2,671 upregulated and 2,476 downregulated genes in the meniscus. In severe OA cartilage, we confirmed the upregulation of genes such as *ADAMTS14* and *COL10A1*, while genes including *ADM* and *GADD45B* were downregulated, consistent with previous studies^16^. In the severe stage meniscus, we further observed significantly increased expression of genes associated with calcification and fibrosis, including *IHH*, *TGFA*, and *COLGALT2* (**Fig. 1b-c, Supplementary Table S2-3**).

Gene Ontology (GO) analysis showed that genes upregulated in severe OA cartilage were enriched for terms such as “Extracellular matrix organization” and “Inflammatory response,” while downregulated genes were enriched for “Focal adhesion PI3K Akt mTOR signaling” (**Supplementary Fig. S2a**). Similarly, in the meniscus, severe OA-upregulated genes were enriched for “Extracellular matrix organization” and “Inflammatory response,” whereas downregulated genes tended to be enriched for terms such as “Fatty acid omega oxidation” (**Supplementary Fig. S2b**).

### Integration of eQTL analysis in cartilage and meniscus with GWAS

Comparison of eQTL effect sizes across different ancestries revealed a shared genetic regulatory architecture, with consistent directionality observed between the European (EUR) population and the American (AMR) and African (AFR) populations (**Supplementary Fig. S3**). Based on this observed correlation in the genetic regulatory architecture, we performed an integrated eQTL analysis on the combined cohort, adjusting for ancestry-driven confounding factors using genotype principal components.

Our integrated analysis identified 1,128 eGenes in cartilage and 1,255 eGenes in the meniscus using the combined data from normal and OA donors. Of these, 692 genes were shared between both tissues, while 436 and 563 genes were specific to cartilage and meniscus, respectively. Colocalization analysis with 1,413 OA GWAS SNVs using the Regulatory Trait Concordance (RTC) method identified 36 candidate targets in cartilage and 40 in the meniscus with an RTC score > 0.9 (Shared: 24, Cartilage-specific: 12, Meniscus-specific: 16) (**Supplementary Table S4**). Additionally, colocalization analysis with *coloc* using All OA and Knee OA GWAS data identified 26 genes in cartilage and 32 in the meniscus with a high posterior probability (PP4 > 0.75) of sharing a causal variant (Shared: 15, Cartilage-specific: 11, Meniscus-specific: 16) (**Supplementary Table S5)**. Moreover, an evaluation of eQTL signals for genes colocalized with OA GWAS using *mashr* confirmed a high degree of sharing in effect signs across sex and disease severity for most genes (**Supplementary Fig. S4, Supplementary Table S6-7**).

Focusing on genes that were both colocalized with OA GWAS signals and identified as differentially expressed genes (DEGs), we extracted five genes in cartilage and three in the meniscus. Among these, genes showing concordance between the direction of expression change in severe OA (vs. normal) and the direction of the eQTL effect driven by the OA risk allele included *CDK2AP1* and *RHCE* in cartilage, and *CDK2AP1* and *CLEC18C* in the meniscus. Conversely, genes showing discordant directions included *SLC13A4*, *ITPRID1*, and *DOT1L* in cartilage, and *CLEC18A* in the meniscus. For instance, *CDK2AP1* expression increased with OA grade in both cartilage and meniscus, and the OA risk allele was associated with a positive eQTL effect. In contrast, in cartilage, *DOT1L* expression was decreased in mild and severe OA compared to normal, but the OA risk allele was associated with increased *DOT1L* expression in normal specimens. In the meniscus, *CLEC18A* expression increased in severe OA, whereas the OA risk allele was associated with decreased expression (**Fig. 2d**). Visualization of eQTL effects and GWAS *p*-values for representative colocalized genes identified *SLC44A2* as a common target in both tissues (*Coloc* PPH4; cartilage: 0.97, meniscus: 0.95), *STIMATE* as a cartilage-specific target (*Coloc* PPH4; cartilage: 0.82, meniscus: 0.030), and *SPATA33* as a meniscus-specific target (*Coloc* PPH4; cartilage: 0.067, meniscus: 0.98) (**Fig. 2e**).

**Fig. 2.**
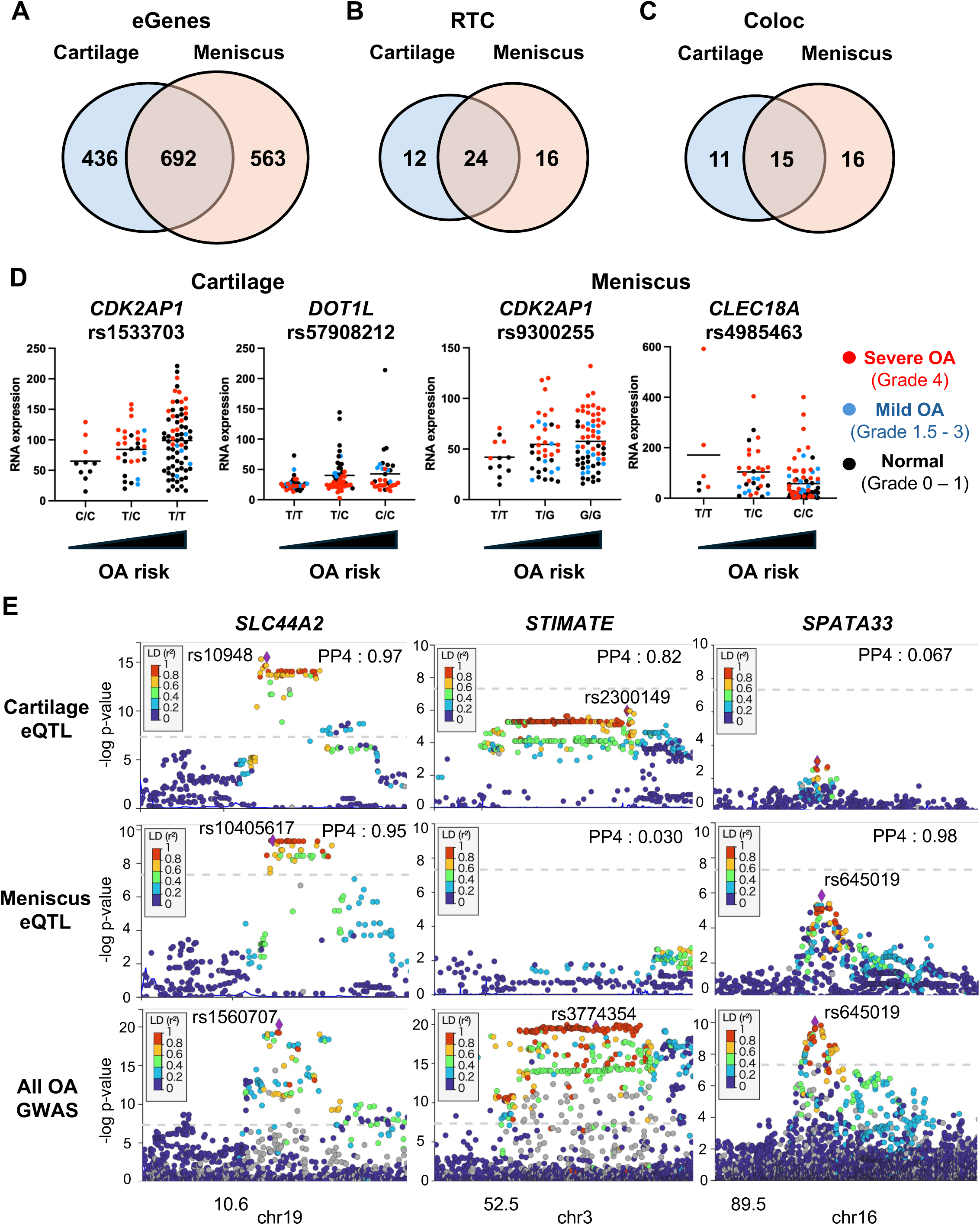
Shared and tissue-specific regulatory effects in cartilage and meniscus. A, Venn diagram showing the overlap of genes with significant *cis*-eQTL effects (eGenes) identified in cartilage (blue) and meniscus (orange). **B,** Colocalization of eQTL signals with OA GWAS loci assessed using the Regulatory Trait Concordance (RTC) method. The bar chart highlights the number of shared and tissue-specific candidate target genes with an RTC score > 0.9. **C,** Colocalization analysis using *coloc* with All OA and Knee OA GWAS datasets (Hatzikotoulas et al., 2025), showing the number of genes with a high posterior probability (PP4 > 0.75) of a shared causal variant. **D,** Representative examples of genes that both colocalize with OA GWAS signals and show differential expression in OA. Box plots display gene expression levels stratified by OA severity (Normal, Mild, Severe) and lead eQTL genotype. *CDK2AP1* (top) shows concordant upregulation by both disease and the risk allele. *DOT1L* (middle) and *CLEC18A* (bottom) illustrate discordant directions, where the risk allele direction opposes the disease-associated expression change. **E,** Regional association plots (LocusZoom-style) for representative loci showing shared (*SLC44A2*) and tissue-specific (*STIMATE* in cartilage, *SPATA33* in meniscus) regulatory architectures. The top panels show eQTL *P*-values, and the bottom panels show GWAS *P*-values. Points are colored by linkage disequilibrium (r^2) with the lead variant.

### Rare variant analysis using Promoter AI

While our preceding analyses of common variants captured genetic risk factors widely shared across the population, OA pathogenesis is also profoundly influenced by rare non-coding variants. Due to their low allele frequencies, these variants are inherently difficult to detect in standard DEG or common eQTL overlaps, yet they can exert substantial effects on individual gene expression. To capture this individualized genetic regulation, we leveraged WGS data to investigate rare regulatory variants. Recent comprehensive evaluations of complex disease heritability have demonstrated that rare variants conferring disease risk are enriched in GWAS loci and tend to exert strong regulatory effects^17–19^.

We therefore integrated our WGS data with a database of predicted impacts on promoter activity for SNVs within ±500 bp of the transcription start site (TSS) using Promoter AI^13^. We specifically focused on the promoter regions of GWAS-proximal genes and eQTL genes colocalizing with GWAS signals. By analyzing SNVs with large absolute Promoter AI scores (≥ 0.2 or ≤-0.2), we evaluated the correlation between the predicted scores and actual gene expression levels in rare variant carriers. The results showed that SNVs with high Promoter AI scores tended to have a larger impact on gene expression in both tissues. A strong correlation between Promoter AI scores and gene expression changes was observed particularly in cartilage (cartilage: *p* = 8.4e-03, meniscus: *p* = 0.090) (**Fig. 3a-d, Supplementary Table S8**). A representative example of this rare-variant-driven regulation is rs36208384 in the *VEGFA* promoter region. Promoter AI predicted a score of-0.26 for this specific variant. Indeed, a significant reduction in *VEGFA* expression was observed exclusively in the rare variant carriers in the meniscus (*p* = 0.032), whereas no clear effect on expression was observed in cartilage (*p* = 0.30), (**Fig. 3e**).

**Fig. 3.**
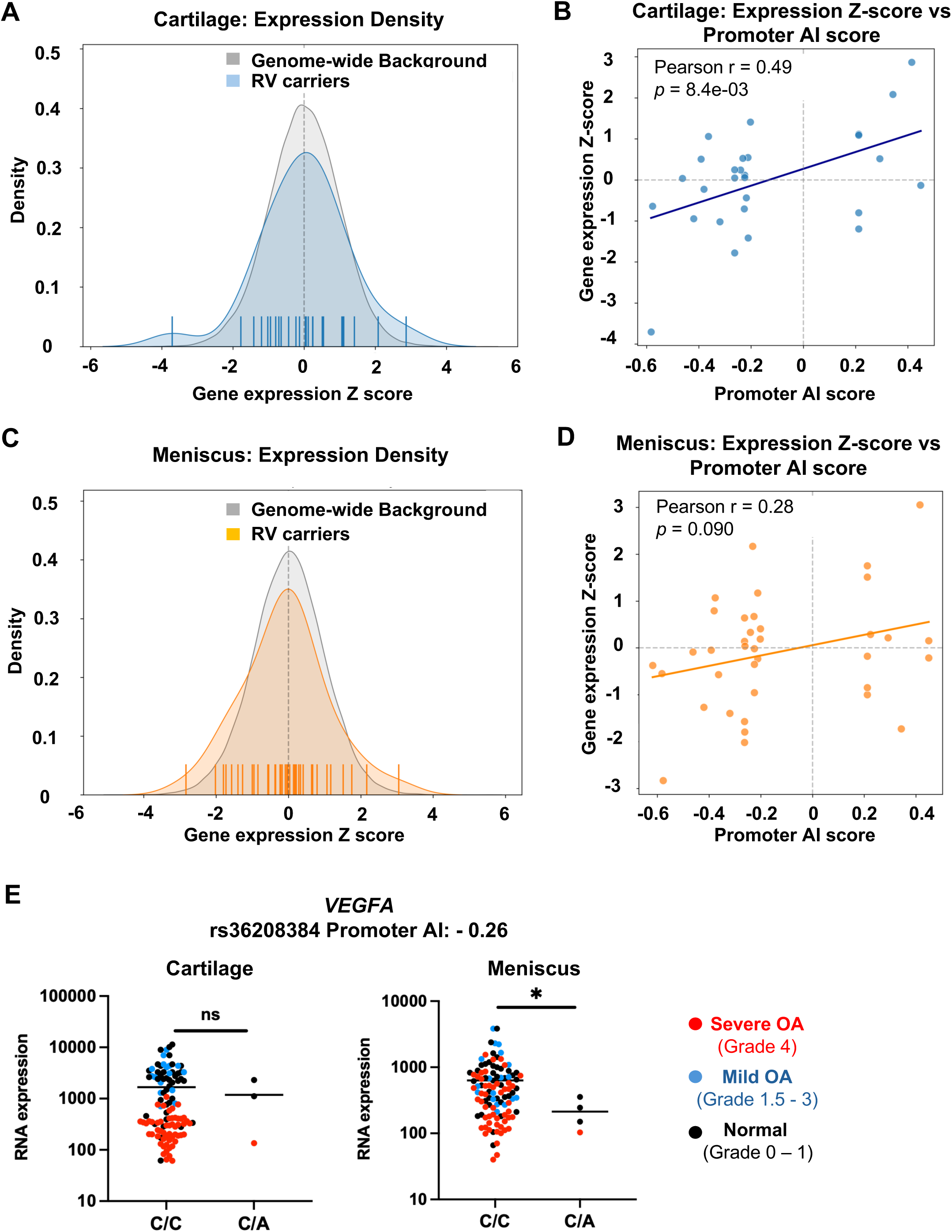
Promoter activity modeling identifies rare regulatory variants with tissue-specific effects. A, Distribution of gene expression Z-scores for carriers of rare variants (RVs) with high predicted promoter impact (Promoter AI score > 0.2 or <-0.2) compared to the genome-wide background in cartilage. **B,** Correlation between predicted promoter activity effects (Promoter AI scores) and observed gene expression Z-scores in RV carriers in cartilage. A significant positive correlation was observed (Pearson *r* = 0.49, *P* = 0.0084). **C,** Distribution of gene expression Z-scores for RV carriers versus genome-wide background in meniscus. **D,** Correlation between Promoter AI scores and observed gene expression Z-scores in meniscus. A similar positive trend was observed, although it did not reach statistical significance (Pearson *r* = 0.28, *P* = 0.090). **E,** Example of a rare promoter variant (rs36208384) in the *VEGFA* locus. The variant has a negative Promoter AI score (-0.26), predicting transcriptional repression. *P*-values were calculated using the Mann-Whitney U test, and the asterisk (*) indicates a *p*-value < 0.05.

### Prediction of cell-type-specific enhancers via Single-nuclei Multiome analysis

Because cartilage and meniscus are composed of multiple cell types, we further sought to identify the cellular origins of the eQTL effects in these tissues. To identify cell-type-specific enhancers in cartilage and meniscus, we performed single-nucleus multiomics analysis on cartilage (56,549 nuclei) and meniscus (34,343 nuclei). We estimated cell-type-specific enhancer regions using SCENT^14^ and attempted to identify disease-associated variants within these regulatory regions by integrating them with OA GWAS SNVs (**Fig. 4a**). Clustering classified cartilage cells into 7 populations (Regulatory chondrocytes (RegC), Reparative chondrocytes (RepC), Homeostatic chondrocytes (HomC), pre-hypertropic chondrocytes (preHTC), pre-fibrochondrocytes (preFC), Fibrochondrocytes (FC), Hypertropic chondrocytes (HTC)) and meniscus cells into 8 populations (HomC, RepC, Immune, FC, preHTC, Endothelial cells, fibrochondrocyte progenitors (FCP), RegC) (**Fig. 4b, Supplementary Fig. S5**). Integrated analysis of SCENT-predicted enhancers and OA GWAS SNVs identified variants potentially involved in enhancer regulation in specific cell types (**Supplementary Table S9**). For example, a peak region containing OA risk SNV rs998584, was predicted to possess enhancer activity for *VEGFA* in the FC (boot *p* = 1.0e-3), FCP (boot *p* = 0.042), Immune cells (boot *p* = 8.0e-5) and preHTC of the meniscus. Interestingly, the peak intensity in this region was downregulated in severe OA samples which coincided with decreased expression of *VEGFA* in FCP. Furthermore, this peak was not detected in articular cartilage, suggesting that it may function as a meniscus-specific enhancer for *VEGFA* (**Fig. 4c**). Notably, referencing the ChIP-Atlas database revealed a STAT3 ChIP-seq peak in the vicinity of rs998584. Coupled with the observation that *STAT3* expression is concurrently downregulated in the FCP of OA samples, these findings implicate STAT3 as a crucial regulator of this enhancer’s activity (**Supplementary Fig. S6**).

**Fig. 4.**
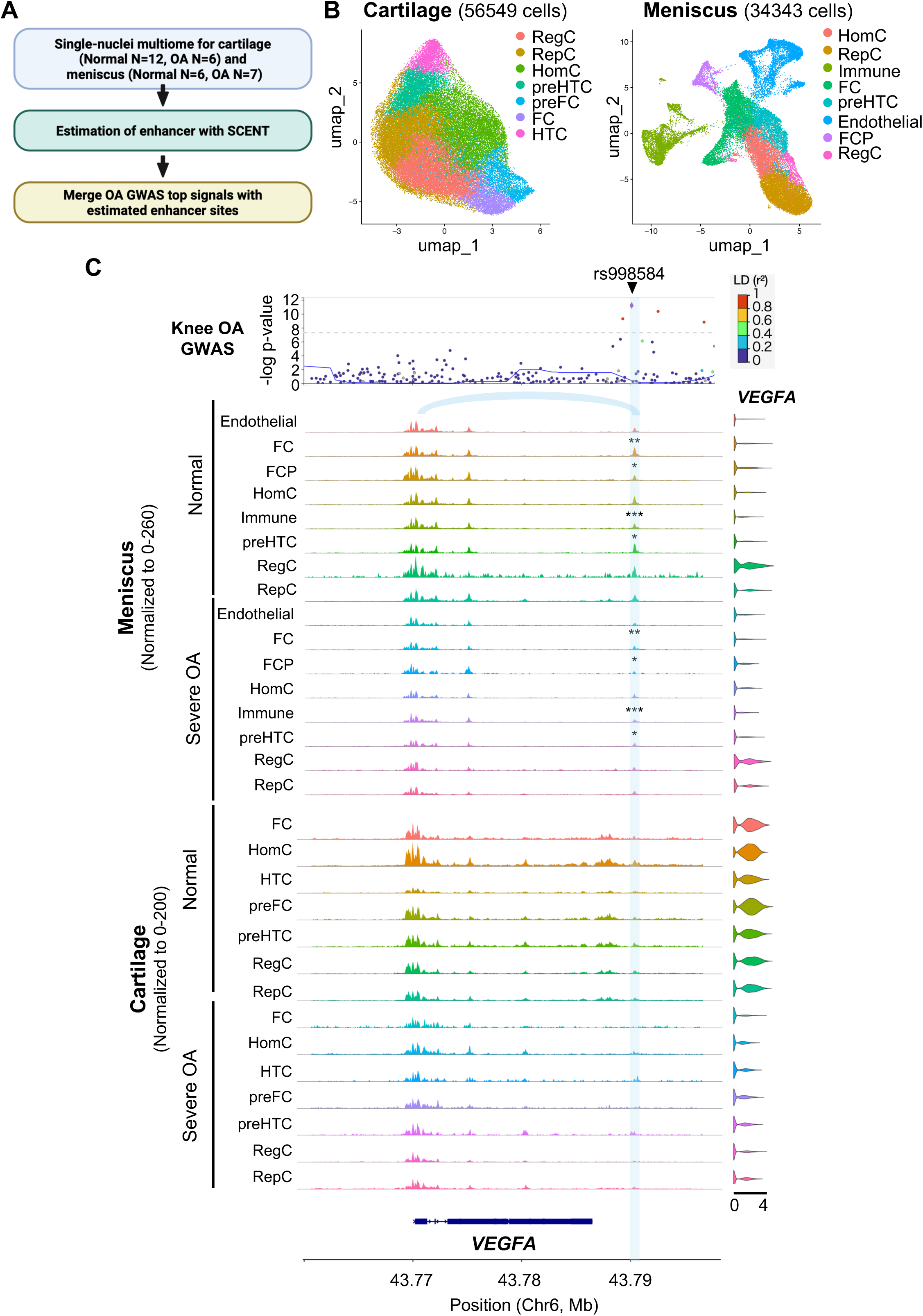
Cell-type–resolved regulatory landscapes in cartilage and meniscus. A, Schematic of the workflow for identifying cell-type–specific enhancers. Single-nuclei multiome data were used to model enhancer-gene links via SCENT, and OA GWAS variants were mapped to these predicted enhancers. **B,** UMAP projections of single-nuclei multiome profiles (WNN graph) from cartilage (left; 56,549 cells) and meniscus (right; 34,343 cells), colored by annotated cell populations. (RegC: Regulatory chondrocytes, RepC: Repair chondrocytes, HomC: Homeostatic chondrocytes, preHTC: pre-Hypertrophic chondrocytes, FC: Fibrochondrocytes, FCP: Fibrochondrocyte progenitors). **C,** Example of a meniscus-specific regulatory element for *VEGFA* containing the Knee OA–associated variant rs998584. Tracks show chromatin accessibility in different cell types for Normal and OA samples. The highlighted region functions as an enhancer specifically in meniscus FCPs and shows increased accessibility in OA conditions. Statistical significance was determined using SCENT boot *p*-values, where *** *p* < 0.005, ** *p* < 0.01, and * *p* < 0.05.

### Cell-type-specific eQTL analysis via Deconvolution

For the deconvolution analysis of our transcriptomic data, we re-analyzed existing single-cell RNA-seq data^20^ for cartilage and meniscus to construct a reference dataset. This dataset (cartilage: 70,411 cells; meniscus: 70,600 cells) was classified into 7 populations for cartilage (RegC, preHTC, preFC, FC, RepC, HomC, HTC) and 8 populations for meniscus (RegC, FC, HomC, preHTC, RepC, FCP, Endothelial, Immune) (**Supplementary Fig. S7-8**). We performed deconvolution analysis on our bulk RNA-seq data using this reference with Bayes Prism^15^. We then re-analyzed eQTL data based on the estimated gene expression levels for each cell population, followed by colocalization analysis (RTC and *coloc*) with the OA GWAS signals (**Fig. 5a**). To comprehensively capture potential causal genes and account for algorithmic differences between the approaches, genes meeting the significance threshold in either method (RTC > 0.9 or coloc PP4 > 0.75) were combined and regarded as colocalized eGenes with OA GWAS. As a result, numerous cell-type-specific eQTL signals colocalizing with OA GWAS were identified in both cartilage and meniscus. For instance, *SLC44A2* showed colocalization with All OA GWAS in both preHTC (*Coloc* PPH4:0.97) and HomC (*Coloc* PPH4:0.83) of the meniscus. In contrast, *SPATA33* showed colocalization in preHTC (Coloc PPH4:0.97) but not in HomC (*Coloc* PPH4:0.074), suggesting cell-type-dependent regulation. Comparing eQTL effects across homologous cell types in cartilage and meniscus, we found that *CLEC18A* was a shared target in preHTC, HomC, and RepC populations in both tissues. Furthermore, six additional genes were shared specifically in RepC. In contrast, cartilage and meniscus-specific eQTL targets were identified, including 21 genes, such as *CDK10* in cartilage RepC, *SEPTIN2* in meniscus RepC and 20 genes, such as *SPATA33*, in meniscus preHTC (**Fig. 5b-d, Supplementary Table S10-11**).

**Fig. 5.**
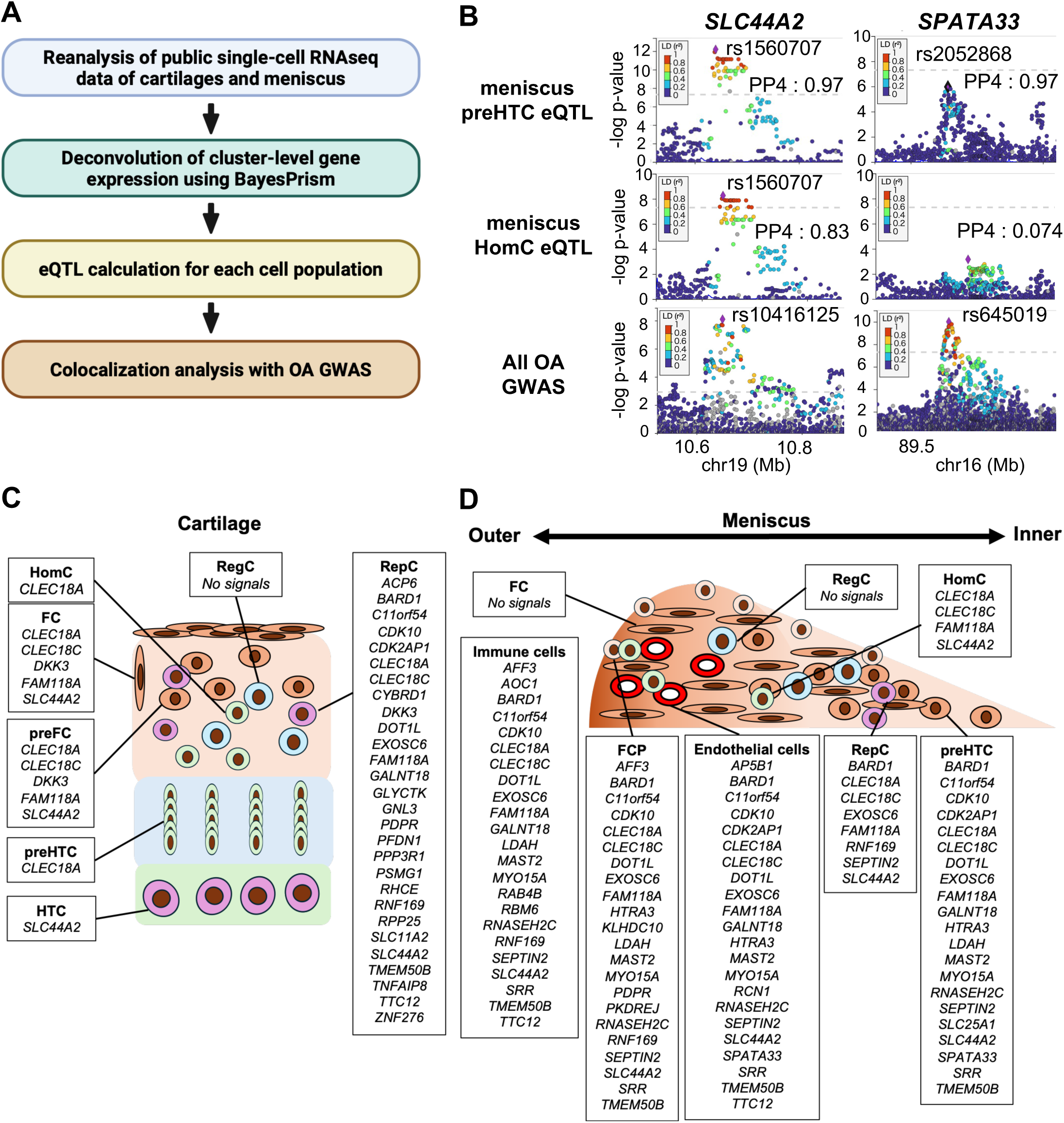
Cell-type–specific regulatory effects revealed by deconvolution-based eQTL mapping. A, Overview of the analysis workflow: reference construction from public single-cell RNA-seq data, deconvolution of bulk RNA-seq using BayesPrism, cell-type–specific eQTL mapping, and colocalization with OA GWAS. **B,** Representative regional association plots of cell-type–specific eQTL signals in the meniscus. *SLC44A2* shows colocalization in both preHTC and HomC populations, whereas *SPATA33* shows preHTC-specific colocalization. **C, D,** Schematic representation of the spatial distribution of cell types in cartilage (**c**) and meniscus (**d**), listing the identified cell-type–specific eGenes co-localized with OA GWAS signals for each population.

### Fine-mapping integrating Deconvolution and Enhancer estimation

For cell-type-specific eQTL signals colocalizing with OA GWAS, we performed fine-mapping using SuSiE, selecting only SNVs within ATAC-seq peaks from the single-nucleus multiome analysis. Here, we improved analysis precision by restricting the analysis to signals present in promoter regions or SCENT-predicted enhancer regions (**Fig. 6a**). Consequently, we successfully identified multiple high-precision eQTL signals supported by multi-omic evidence in both cartilage and meniscus (**Supplementary Table S12-13**). As a specific example, rs4985407 in meniscal RepC showed an eQTL effect where the OA risk allele reduced the expression of both *CLEC18A* and *EXOSC6* (**Fig. 6b**). Although rs4985407 is located in the *EXOSC6* promoter region, SCENT analysis estimated that it is also contained within a region possessing enhancer activity for *CLEC18A* (boot *p* = 0.014). Furthermore, the ATAC peak in this region and *CLEC18A* expression in RepC were increased in OA specimens compared to normal controls, suggesting the existence of a disease-specific regulatory mechanism (**Fig. 6c**). Referencing the ChIP-Atlas database revealed a ETS1 ChIP-seq peak in the vicinity of rs4985470. Coupled with the observation that *ETS1* expression is concurrently upregulated in the RepC of OA samples, these findings implicate ETS1 as a crucial upstream regulator of this enhancer’s activity (**Supplementary Fig. S9**).

**Fig. 6.**
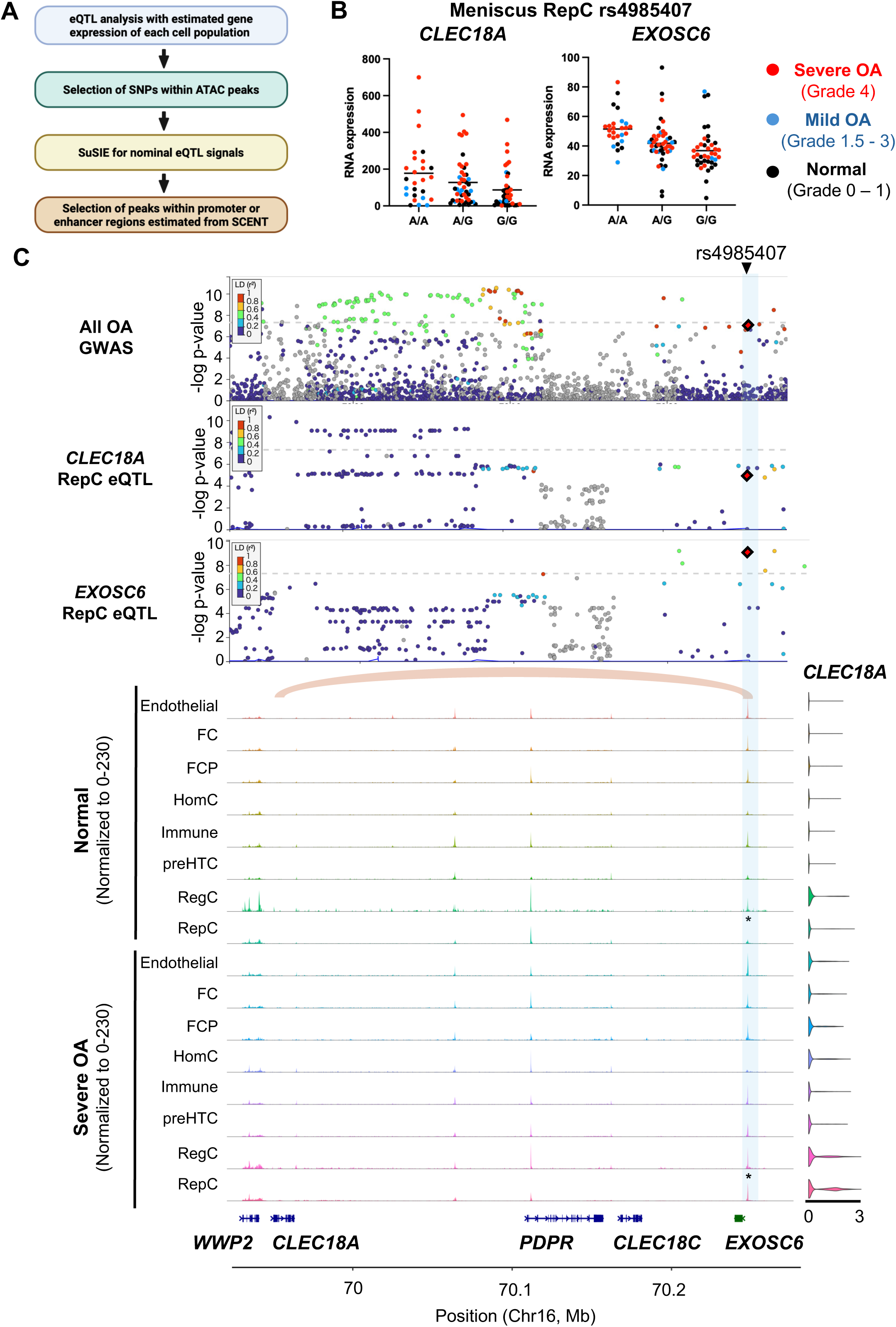
Fine-mapping of regulatory variants integrating chromatin accessibility and enhancer annotations. A, Schematic of the fine-mapping strategy using SuSiE. Candidate variants were restricted to those located within single-nuclei ATAC-seq peaks and further prioritized if they fell within SCENT-inferred regulatory regions (enhancers or promoters). **B,** Fine-mapped locus at rs4985407 showing eQTL effects on *CLEC18A* and *EXOSC6* in meniscus RepC cells. **C,** Chromatin accessibility tracks at the *CLEC18A/EXOSC6* locus in meniscus RepC cells. Accessibility is increased in OA samples compared to Normal, suggesting disease-responsive regulatory remodeling that is disrupted by the risk variant. Statistical significance was determined using SCENT boot *p*-values (* *p* < 0.05).

### Drug response prediction targeting shared pathogenic signatures in cartilage and meniscus

Finally, to explore potential therapeutic strategies, we performed drug response prediction using the SigCom LINCS database^21^, targeting the shared regulatory and transcriptomic signatures identified in both cartilage and meniscus. The analysis utilized input gene sets representing these shared mechanisms, consisting of eGenes colocalizing with OA GWAS signals and DEGs showing consistent expression changes in severe OA across both tissues. We identified four candidate compounds capable of reversing these shared pathogenic profiles. Withaferin-a, TW-37, MG-132, and Dexamethasone consistently demonstrated strong negative connectivity scores across multiple cell lines for both the genetically-driven and disease-associated signatures (**Table1, Supplementary Table S14**).

## Discussion

OA has traditionally been characterized as a degenerative disorder primarily affecting articular cartilage. Consequently, most genetic and transcriptomic studies to date have focused on cartilage as the primary tissue of interest^22^. However, recent clinical and epidemiological evidence indicates that OA should be conceptualized as a “whole-joint” disease involving the meniscus and synovium, rather than a condition confined solely to cartilage^4,23^. In particular, meniscal damage and degeneration are established as major risk factors for the development of knee OA^12,24^. Despite its clinical significance, the genetic basis of gene expression regulation in the meniscus and its contribution to OA susceptibility have remained largely unexplored from a molecular genetic perspective.

A crucial strength of this study lies in the inclusion of high-quality normal tissues, and from knees with mild OA distinguishing it from previous studies that largely analyzed surgical specimens from end-stage OA patients undergoing total joint replacement. This prior reliance on end-stage tissues limits the ability to evaluate baseline genetically determined regulation in the normal state and during early stages of disease. By utilizing normal cartilage and meniscus samples, we identified eQTL signals for genes whose expression is substantially downregulated or lost during disease progression.

Integration of DEGs from transcriptomic analysis with eQTL genes colocalizing with OA GWAS signals identified five genes in cartilage and three in the meniscus. These are considered key genes that either contribute to OA pathogenesis or reflect disease progression. Among them, *CDK2AP1* was identified in both tissues as a gene upregulated in OA compared to normal specimens and positively regulated by OA risk alleles. *CDK2AP1* is known to inhibit DNA replication and arrest cell proliferation by inhibiting Cyclin-dependent kinase 2 (CDK2) activity^25^. The dual findings of upregulation in OA specimens and upregulation by genetic risk suggest that cell cycle arrest caused by *CDK2AP1* overexpression is both a cause and a consequence of OA pathology, suggesting that *CDK2AP1* represents a promising therapeutic candidate targeting both tissues.

In contrast, we observed discordance between expression changes and eQTL effects, such as *DOT1L* in cartilage. Marigorta et al. proposed that when genetic regulation aligns with tissue expression changes, the gene is a likely causal driver, whereas discordance suggests a response or compensatory mechanism^26^. *DOT1L* is a cartilage-protective factor that suppresses Wnt signaling via histone methylation^27^. Its downregulation in OA cartilage implies a breakdown of this protective mechanism^28^; paradoxically, the risk allele correlates with higher baseline expression in health. This indicates that strict homeostasis is essential; constitutively elevated expression may restrict regulatory flexibility, constraining adaptive responses under disease-associated stress. Consequently, our eQTL data cautions that simple functional supplementation of *DOT1L* does not guarantee therapeutic efficacy. Thus, integrating DEG and eQTL directionality disentangles causal drivers from context-dependent consequences within pathogenic networks.

Furthermore, by leveraging WGS data, this study focused on the functional significance of rare variants. We introduced the latest deep learning model, Promoter AI^13^, to predict the function of observed rare variants and enable their integration with actual transcriptomic data. At the individual gene level, important insights symbolizing the utility of this approach were obtained. A representative example is a variant in the *VEGFA* promoter region (rs36208384). Promoter AI predicted a strong suppressive effect on expression for this variant, and carriers in the meniscus showed significantly lower *VEGFA* expression compared to non-carrier.

These results demonstrate that combining WGS with deep learning prediction is a powerful tool for rare variants near GWAS SNVs, and for uncovering tissue-specific pathogenic mechanisms.

Intriguingly, our enhancer prediction using SCENT further identified a meniscus-specific enhancer region for *VEGFA* harboring rs998584, a SNV previously associated with knee OA particularly within FCPs. Notably, in severe OA samples, the chromatin accessibility at this enhancer was markedly diminished, whereas no corresponding peak was observed in cartilage. Taken together with the Promoter AI predictions, these findings suggest that a specific genomic background leads to the downregulation of *VEGFA* in the meniscus, thereby predisposing the joint to OA. The meniscus is characterized by a distinct vascular architecture, with a well-perfused outer zone and an avascular inner zone^29^. Given that FCPs are predominantly distributed in the superficial layers of the outer zone^30^, our results imply that maintaining *VEGFA* expression is a vital homeostatic mechanism unique to the vascularized regions of the meniscus. Consequently, therapeutic strategies aimed at supplementing *VEGFA* expression specifically within the meniscus may represent a promising avenue for OA intervention.

Another notable aspect of this study is the elucidation of cell-type-specific gene regulatory mechanisms using public single cell transcriptomic data^20^ and deconvolution techniques with BayesPrism^15^. The meniscus is a structurally and cytologically highly heterogeneous tissue composed of an avascular, cartilage-like inner zone and a vascular, fibrocartilaginous outer zone^30^. In such a tissue with mixed cell populations, conventional bulk analysis carries a risk that variations in cell composition due to sampling sites act as noise, masking cell-intrinsic gene regulatory changes. Therefore, using existing single-cell RNA-seq data as a reference, we successfully distinguished genes regulated commonly across multiple cell types from genes showing eQTL effects only in specific cell types via Bayes Prism-based deconvolution. For example, we identified *CLEC18A* as a shared regulatory target across multiple cell types (HomC, RepC, preHTC) in both cartilage and meniscus, highlighting it as a molecule of common importance in OA pathology across joint tissues. In contrast, we also identified tissue-specific eQTLs, such as *SEPTIN2* in meniscus RepC and *SPATA33* in meniscus preHTC. Despite the transcriptomic similarity of these cells to their cartilage counterparts, these findings suggest that these molecules may function specifically within the meniscal microenvironment. This distinction implies that OA susceptibility involves both shared molecular drivers and tissue-specific mechanisms unique to the meniscus. The cell-type-resolved comparative analysis provides critical insights into how genetic risks manifest in homologous cell populations residing in distinct tissues.

Moreover, the approach integrating ATAC-seq peak information based on single-nucleus multiomics with eQTL analysis for the functional interpretation of OA GWAS signals is groundbreaking in this field. We employed a strategy of limiting variants for fine-mapping via SuSiE to SNVs located within open chromatin regions in promoters or SCENT-predicted enhancers. Using tissue-and cell-specific chromatin accessibility as a biological prior filter reduced false positives and enabled more accurate identification of causal variants. To demonstrate the efficacy of this approach, we focused on the *CLEC18A/EXOSC6* locus.

ATAC-seq data showed specifically increased chromatin accessibility in this region in severe OA meniscal RepC, likely reflecting an epigenetic response to induce *CLEC18A* expression under disease conditions. However, the risk allele was found to inhibit this enhancer activity, resulting in reduced *CLEC18A* expression. *CLEC18A* is known as a C-type lectin that binds to polysaccharides in a Ca2+-dependent manner^31^. The fact that *CLEC18A* expression and chromatin accessibility were specifically elevated in meniscal RepC in this study suggests a potential involvement of this molecule in tissue repair or protective processes under OA pathology.

Finally, as an exploratory step toward clinical translation, we conducted an *in silico* drug discovery search using SigCom LINCS^21^ to identify compounds that have the potential to counteract the eQTL-mediated effects and reverse the disease-associated transcriptomic signatures. This analysis yielded four candidate drugs demonstrating significant potential as therapeutic agents for OA by simultaneously targeting both the meniscus and cartilage.

Notably, dexamethasone is already utilized clinically as an intra-articular injection for the treatment of OA^32^. Furthermore, analysis showed the possibility for therapeutic use against OA for TW-37^33^, MG-132^34^, and withaferin A^35^.

In the future, integrating three-dimensional genome structural analyses such as Hi-C is expected to enable direct verification of physical interactions between eQTLs and enhancers. Such high-resolution data are essential for improving the accuracy of next-generation regulatory variant effect prediction models like AlphaGenome^36^. To this end, the systematic accumulation of primary human data from normal and diseased states in each tissue constituting the locomotor system is crucial for the advancement of OA research.

In conclusion, this study brings a new dimension to elucidating the genetic basis of OA by covering cartilage and meniscus, thereby integrating chromatin structure and gene expression regulation at the cell-type level. In particular, the identification of unique gene regulatory mechanisms in the meniscus and cell-type-specific eQTLs provides robust molecular evidence for redefining OA as organ failure of the entire joint.

## Materials and Methods

### Ethical approvement

Human tissue collection was approved by the Scripps Human Subjects Committee (20-7636).

### Knee Tissue Procurement

We have established a systematic approach for procurement and processing of intact human knee joints from donors aged 20–80 through a long-standing collaboration with Lifesharing, a local Organ Procurement Organization (OPO). Donors are selected based on predefined criteria to ensure high-quality, unbiased data while minimizing variability. Exclusion criteria include recent knee injury or surgery, autoimmune disease, inflammatory arthritis, prolonged immobilization, neuromuscular disorders, chronic medication use, metabolic disease (osteoporosis, diabetes), sepsis, chemotherapy, and other severe comorbidities. Following our standardized protocol, both knees are resected 10 cm above and below the joint line with skin and capsule intact, and shipped on wet ice to the laboratory within 24 hours for immediate processing. The OPO also collects peripheral blood and a standardized medical history, including demographics, trauma, comorbidities, medication use, and prior OA treatments, providing essential context for downstream multiomic analyses^37^. As an additional source, tissues are obtained from OA patients undergoing TKA at Scripps Clinic via an established collaboration with an orthopedic surgeon. For all OA donors, radiographs and Kellgren/Lawrence scores are available, along with detailed clinical information (medications, comorbidities, pain treatments, radiographs, MRI and joint involvement based on ACR criteria). A central database integrates donor history, clinical data, grading results, and the location of paraffin-embedded blocks, frozen tissues, cells, and extracts (nuclei, RNA, and DNA).

### Knee Tissue Resection

Upon opening the joint capsule, high-resolution photographs were taken of all cartilage areas and other tissues. All cartilage surfaces were graded macroscopically using the Outerbridge system, and a total knee score is calculated. We collected menisci samples spanning the central weight-bearing part of each condyle and the central area of the entire tibial plateau and the entire IPFP.

### Tissue Collection and Processing

To minimize tissue degradation, collection and processing was performed using established protocols^38,39^. For omics analyses, we isolated nucleic acids directly from tissues and nuclei for single-nucleus multiome.

### Single-nuclei Analysis

We used the 10x Genomics Chromium Controller, targeting 10,000 cells per sample for high-quality analyses. The library preparation was performed following manufacturer’s protocols; an optimized nuclei isolation buffer was employed for nuclear isolation. For library preparation, ATAC-seq transposition was performed at 37°C for 30 minutes, followed by PCR amplification with optimized cycle numbers. For samples intended to analyze responses to mechanical loading, conditions were adjusted to sensitively detect chromatin structural changes. Sequencing depth was set to 5 million reads per cell to achieve high-resolution transcript profiling^40^.

### Sequencing protocols

RNA-seq libraries were prepared using the NEBNext Ultra II Directional Prep Kit (NEB) and sequenced on an Illumina NovaSeq with 150-bp paired ends. For whole-genome sequencing (WGS), libraries were prepared from gDNA using the TruSeq DNA PCR-Free Library Preparation Kit (Illumina), adjusted to an average insert size of approximately 350 bp, and sequenced on the Illumina platform (NovaSeq X Plus) with 2×150-bp paired-end reads. For single-nuclei multiome analysis, nuclei were isolated from frozen tissues using the Singulator, and libraries were prepared with the Chromium Next GEM Single Cell Multiome ATAC + Gene Expression Reagent Bundle targeting a recovery of 10,000 cells.

### Whole genome sequencing analysis

Low-quality bases and adapter sequences were removed from raw FASTQ files using fastp. The cleaned reads were aligned to the GRCh38 reference genome (excluding ALT, HLA, and decoy contigs) using BWA-MEM^41^. The aligned BAM files were sorted by coordinate using SAMtools^42^, and duplicate reads were identified and marked using GATK MarkDuplicates (v4.5.0.0). To improve the accuracy of variant calling, we applied Base Quality Score Recalibration (BQSR) using the GATK BaseRecalibrator and ApplyBQSR tools, referencing known variants from dbSNP (v138) and the 1000 Genomes Project. Genomic variant call format (gVCF) files were generated for each individual using GATK HaplotypeCaller with the--emit-ref-confidence GVCF option.

### Joint Genotyping and Variant Filtering

Individual gVCF files were consolidated into a GenomicsDB using GATK GenomicsDBImport, and joint genotyping was performed across the cohort using GATK GenotypeGVCFs to generate a raw multi-sample VCF file. The resulting variants were concatenated across chromosomes using GATK MergeVcfs. To obtain high-quality variants, we applied Variant Quality Score Recalibration (VQSR) separately for SNPs and insertions/deletions (indels). For SNPs, the recalibration model was trained using HapMap 3.3, 1000 Genomes Omni 2.5, and 1000 Genomes Phase 1 datasets, with dbSNP (v138) as reference. The annotations used for modeling included QD, MQ, MQRankSum, ReadPosRankSum, FS, SOR, and DP. For indels, the model was trained using the Mills and 1000 Genomes gold-standard set and dbSNP, utilizing QD, DP, FS, SOR, ReadPosRankSum, and MQRankSum as annotations. Finally, the recalibrated SNP variants were annotated with reference rsIDs (dbSNP) using SnpSift.

### RNA Sequencing and Data Processing

Sequencing libraries were prepared and sequenced as 150-bp paired-end reads. Raw FASTQ files were processed using a standardized pipeline based on the GTEx Project workflow^43^. Briefly, adapter trimming and quality control were performed using fastp (v0.23.2) with the --detect_adapter_for_pe option. The trimmed reads were aligned to the GRCh38 human reference genome (noALT_noHLA_noDecoy_ERCC) using STAR (v2.7.10a)^44^.

Gene-level quantification was performed with RSEM (v1.3.1) using the GENCODE v39 annotation. Post-alignment quality control, including the calculation of mapping metrics and transcript coverage, was conducted using RNA-SeQC (v2.4.2). Duplicate reads were identified and marked using MarkDuplicates (Picard tools).

### Differential Expression and Functional Enrichment Analysis

To identify transcriptomic changes associated with disease severity, we performed differential expression analysis comparing mild OA and severe OA to normal tissue samples. DEGs were defined as those meeting the following criteria: a Benjamini-Hochberg adjusted *p*-value < 0.05 and an absolute log2 fold change > 1.

Subsequently, to characterize the biological functions associated with these DEGs, we performed pathway and process enrichment analysis using Metascape (https://metascape.org). The lists of upregulated and downregulated genes were analyzed separately to capture direction-specific biological alterations. We utilized the Gene Ontology (GO) Biological Processes, KEGG Pathway, and Reactome Gene Sets databases.

### Data Preprocessing for eQTL Analysis

Gene expression quantification was performed using RSEM, and count matrices were subset to samples with matching whole-genome sequencing (WGS) data. Gene expression values were normalized using the Trimmed Mean of M-values (TMM) method to account for library size differences. The normalized expression data were formatted for QTL analysis using gene annotations from GENCODE version 39. For genotype data, WGS variants were filtered to include only those present in the corresponding RNA-seq samples. We applied strict quality control filters, retaining variants with a Variant Quality Score Log-Odds (VQSLOD) ≥0 and a MAF > 0.05. To ensure high variant quality, we further restricted the analysis to variants intersecting with the 1000 Genomes Project Phase 3 reference panel (GRCh38). To correct for population stratification, we performed a PCA on the filtered genotype data using QTLtools. The top five genotype PCs were extracted and used as covariates in the subsequent analysis. *cis*-eQTL mapping was performed using QTLtools cis mode.

Nominal p-values were calculated for all variant-gene pairs within a 1 Mb window upstream and downstream of the TSS. To correct for multiple testing, we applied permutation testing with 1,000 permutations (--permute 1000) to determine the significance of the associations.To correct for multiple testing across genes, we calculated the False Discovery Rate (FDR) using the QTLtools^45^ standard procedure. Genes with an FDR < 0.05 were defined as significant eGenes.

### Colocalization Analysis using Regulatory Trait Concordance (RTC)

To assess whether the identified *cis*-eQTLs share the same causal signal with OA risk variants, we calculated the RTC score using QTLtools. We compiled a comprehensive list of OA-associated variants from the GWAS Catalog (trait: Osteoarthritis) and all significant SNVs identified in a recent large-scale meta-analysis^7^. For each significant eQTL (eGene), we computed the RTC^46^ score against these GWAS variants, accounting for local LD structure using the hg38 recombination hotspot map. The top five genotype principal components were included as covariates to correct for population structure. An RTC score > 0.9 was considered indicative of a shared causal variant between the eQTL and the GWAS trait.

### Colocalization Analysis with Coloc

To investigate whether the identified OA-associated variants share a common causal variant with the *cis*-eQTLs in cartilage/meniscus tissues, we performed Bayesian colocalization analysis using the coloc R package^47^. We utilized the coloc.abf function, which computes approximate Bayes factors (ABF) for five hypotheses, where Hypothesis 4 (H4) represents the existence of a shared causal variant. Summary statistics (beta coefficients, standard errors, and *P*-values) from the OA GWAS meta-analysis^7^, and our tissue-specific eQTL analysis were merged based on rsIDs. For the eQTL dataset, SE were estimated from the beta coefficients and *P*-values. We considered genes with a posterior probability for PP4 > 0.75 as having strong evidence for colocalization.

### Comparative analysis with Mashr

To characterize the sharing and specificity of genetic regulatory effects across different biological contexts—specifically disease states (Normal vs. Mild OA vs. Severe OA), and sexes (Male vs. Female)—we applied the Multivariate Adaptive Shrinkage (mashr) framework^48^. To prevent ascertainment bias favoring a specific condition, we employed a “union of lead variants” strategy for variant selection. First, lead *cis*-eQTLs were identified independently within each subgroup. These candidates were pooled, and for each gene, a single representative variant was selected based on the minimum nominal p-value across all compared subgroups. Since SE were not directly available in all summary statistics, we estimated the SE from the beta coefficients and nominal p-values. We constructed a strong dataset of these selected effects and a random dataset of null effects sampled from non-significant variants to estimate the residual correlation structure. The mash model was fitted using canonical covariance matrices to capture broad patterns of heterogeneity. Posterior effects were evaluated using the Local False Sign Rate (lfsr); eQTLs were defined as “shared” if they showed lfsr < 0.05 and consistent effect directions across conditions, and “context-specific” if they showed lfsr < 0.05 in only a subset of conditions.

### Rare Variant Analysis and Promoter Activity Prediction

To investigate the impact of rare variants on gene expression in OA-relevant loci, we extracted high-quality SNVs (VQSLOD ≥ 0) located within promoter regions, defined as < 500 bp around the canonical TSS based on GENCODE v39 annotations. We filtered for rare variants defined as having a MAF < 0.05 in our local cohort. To ensure rarity in the general population, we further excluded variants with an MAF > 0.05 in the 1000 Genomes Project Phase 3 dataset.

We focused on RVs located in the promoters of (1) genes located near OA GWAS risk loci and (2) eGenes identified as colocalizing with OA GWAS signals in our *cis*-eQTL analysis. The functional impact of these RVs on transcriptional activity was predicted using PromoterAI^13^. We prioritized variants with a predicted substantial effect on promoter activity, defined as a PromoterAI score <-0.2 or > 0.2.

Finally, to validate these predictions, we examined the relationship between the predicted PromoterAI scores, and the actual gene expression levels derived from our bulk RNA-seq data. For carriers of these rare variants, we compared the predicted scores with TMM-normalized gene expression values using Pearson correlation analysis.

### Single-nuclei Multiome Analysis and Functional Annotation Data Preprocessing and Clustering

Raw files were demultiplexed and aligned to the human reference genome (GRCh38) using the Cell Ranger ARC pipeline. The resulting filtered feature-barcode matrices were loaded into R using Seurat^49^ and Signac^50^. For the RNA assay, gene expression counts were normalized using SCTransform. For the ATAC assay, term frequency-inverse document frequency (TF-IDF) normalization and singular value decomposition (SVD) were performed. We integrated the two modalities using the Weighted Nearest Neighbor (WNN) method to derive a joint cell-specific representation, which was used for graph-based clustering and cell type annotation.

### SCENT-based Enhancer Modeling

We utilized SCENT^14^ to predict functional enhancer-gene links. For each cell type, SCENT applies a Poisson regression model to associate chromatin accessibility at peak regions with the expression of nearby genes, accounting for technical noise. Significant links were determined based on a bootstrapped *p*-value < 0.05. We compiled 1561 OA susceptibility variants from the GWAS Catalog and the summary statistics^7^. We then mapped these variants to the genomic coordinates of the SCENT-predicted enhancers. This allowed us to identify specific enhancers in cartilage and meniscus cell subpopulations that are likely to mediate OA genetic risk.

### Deconvolution and Cell-type Specific eQTL Analysis

To dissect cell-type specific regulatory effects from bulk tissue data, we performed computational deconvolution using BayesPrism^15^. We utilized a publicly available single-cell RNA-seq dataset of human cartilage and meniscus^20^ as a reference to define cell-type gene expression signatures. Using the bulk RNA-seq count matrices and the single-cell reference, we estimated the cell-type specific gene expression levels for each sample. The inferred expression values were scaled to a library size of 10^5^ to facilitate comparison across samples. Subsequently, these estimated cell-type specific expression matrices were used as input for *cis*-eQTL mapping. The downstream data processing workflow—including TMM normalization, covariate correction using genotype PCs, and permutation testing with QTLtools—was identical to that described above for the bulk eQTL analysis.

### Fine-mapping and Functional Annotation of Cell-type Specific eQTLs

To prioritize putative causal variants within the identified cell-type specific eQTL signals co-localized with OA GWAS, we performed statistical fine-mapping integrated with chromatin accessibility data. Initially, to restrict the analysis to biologically relevant genomic regions, we filtered for variants located within ATAC-seq peaks identified by the Cell Ranger ARC pipeline.

It should be noted that these peaks represent aggregate chromatin accessibility across all single nuclei. Using this filtered variant set, we computed the LD correlation matrix for the variants using PLINK 1.9. We then applied the SuSiE model to the cell-type specific nominal summary statistics and the estimated LD matrix to calculate the 95% credible sets of variants likely containing the causal effect. Subsequently, to link these statistical candidates to specific regulatory mechanisms, we annotated the variants within the credible sets based on their overlap with functional elements. We prioritized variants located in promoter regions, defined as ±2 kb from the TSS, or in cell-type specific enhancers predicted by SCENT with a significance threshold of a bootstrapped *P*-value < 0.05. This integrative approach enabled the identification of high-confidence causal variants supported by both statistical fine-mapping and cell-type specific regulatory evidence.

### SigCom LINCS

To identify potential therapeutic compounds capable of reversing the observed disease signatures, we performed an *in silico* drug repurposing analysis using the SigCom LINCS^21^ platform. As input, we utilized the directional expression changes of DEGs alongside eGenes that colocalized with GWAS risk alleles. Specifically, this signature reversal search was systematically conducted across six distinct target categories: DEGs specific to the cartilage, specific to the meniscus, and shared across both tissues, as well as eGenes colocalized with GWAS loci that are specific to the cartilage, meniscus, and shared across both tissues.

## Statistical Analysis

All statistical analyses were performed using R (version 4.5.1). Comparison of effect sizes between groups was performed using the Wilcoxon rank-sum test and Kolmogorov-Smirnov (KS) test. Unless otherwise specified, a *p*-value < 0.05 was considered statistically significant.

## Data availability

The raw sequencing data generated in this study have been deposited in the dbGAP database and will be made publicly available upon acceptance of the manuscript. Publicly available datasets analyzed in this study are available from their respective repositories: single-cell RNA-seq data from Swahn et al. (2022) and OA GWAS summary statistics from Hatzikotoulas et al. (2025)^7^.

## Data Availability

The raw sequencing data generated in this study have been deposited in the dbGAP database and will be made publicly available upon acceptance of the manuscript. Publicly available datasets analyzed in this study are available from their respective repositories: single-cell RNA-seq data from Swahn et al. (2022) and OA GWAS summary statistics from Hatzikotoulas et al. (2025).

## Acknowledgements

The authors thank Shiho Makino, Shohei Takihira, and all the staff of the Department of Systems Biomedicine at Institute of Science Tokyo for their support and advice. M. Payad, Z. Flores, and the team at Lifesharing, San Diego provided access to the precious gift of tissue donations from our local San and Imperial County communities that made our research possible. W. Bugbee, D. D’Lima, and N. Glembotski supported procurement of joint tissues from patients undergoing knee arthroplasty. Computations were partially performed on the NIG supercomputer at ROIS National Institute of Genetics. The schematics in Fig.1a and the graphical abstract were created with BioRender (biorender.com) under a publication license.

This work was supported by Japan Society for the Promotion of Science (JSPS) KAKENHI (Grant Numbers JP20H05696), Japan Agency for Medical Research and Development (AMED) (Grant Numbers JP22gm0010009, JP24jf0126010, JP24gm2010002), and National Institutes of Health (NIH) (Grant Number R01AR080127) to H.A., and R01AG049617 and UC2 AR082186 to ML.

## Author contributions

Conceptualization: Y.U., Y.F., Y.K., M.L., and H.A.; methodology: Y.U., Y.F., H.S., M.U., M.O., R.N., Y.K., M.L., and H.A.; validation: Y.U., H.S., T.C., T.M., M.O., Y.N.; formal analysis: Y.U., Y.F., and H.S.; investigation: Y.U., Y.F.,T.C., T.M., M.O., Y.N.; resources: Y.F., H.S., M.O., K.T., M.L., and H.A.; data curation: Y.U, H.S., M.O., Y.N..; writing – original draft preparation: Y.U., F.Y.,; writing – review and editing: Y.U., Y.F., H.S., M.U., T.C., T.M., R.N., M.L., Y.K., and H.A.; visualization: Y.U., M.U., R.N.; supervision: K.T., M.L., Y.K. and H.A.; project administration: H.A.; funding acquisition: H.A.

---

**Table1: Candidate therapeutic compounds for the reversal of disease-associated eQTLs and transcriptomic signatures**

This table lists the candidate compounds identified to simultaneously reverse the directional expression changes of DEGs shared between the articular cartilage and meniscus, as well as the transcriptomic signatures of shared eGenes colocalized with GWAS risk alleles. P-values were calculated using the SigCom LINCS platform to evaluate the significance of signature reversal.

## Supplementary Figures

**Supplementary Fig. S1 | Genetic ancestry composition across all samples.**

**A**, PCA plot of genotype data illustrating population structure.

**B**, Pie chart showing the distribution of inferred genetic ancestries.

**Supplementary Fig. S2 | Functional enrichment analysis of differentially expressed genes.**

**A, B,** Bar plots showing the top enriched Gene Ontology (GO) and pathway terms for genes upregulated (top) and downregulated (bottom) in severe OA compared to normal tissue in cartilage (**a**) and meniscus (**b**). Bars are colored by-log10(*P*-value).

**Supplementary Fig. S3 | Consistency of eQTL effect sizes across ancestries.**

**A**, Scatter plots comparing eQTL effect sizes (Beta) in cartilage between the European (EUR) and American (AMR) populations (left), and between the EUR and African (AFR) populations (right).

**B**, Scatter plots comparing eQTL effect sizes in meniscus between the EUR and AMR populations (left), and between the EUR and AFR populations (right).

**Supplementary Fig. S4 | Comparison of eQTL effect sizes across sex, and disease severity.**

**A,** Comparison of eQTL beta distributions between male and female samples in cartilage and meniscus.

**B,** Comparison of eQTL beta distributions between Normal and Severe OA samples. A shift toward larger effect sizes in Severe OA is observed in cartilage (*P* = 4.2e-10, KS test).

**Supplemtary Fig. S5 | Cell type annotation of the single-nuclei multiome cartilage dataset.**

**A,** UMAP feature plots showing the expression of canonical marker genes used to annotate cell clusters in the cartilage single-nuclei multiome dataset.

**B,** Feature plots showing marker gene expression across the defined clusters in the dataset.

**Supplementary Fig. S6 | Epigenetic binding and expression profile of STAT3.**

**A**, Visualization of STAT3 ChIP-seq peaks at the *VEGFA* genomic locus, retrieved from the ChIP-Atlas database.

**B**, Single-nucleus gene expression levels of *STAT3* within the meniscus, derived from snMultiome sequencing data.

**Supplementary Fig. S7 | Construction of the single-cell RNA-seq reference of cartilage for deconvolution.**

**A,** UMAP visualization of the re-analyzed public single-cell RNA-seq dataset of cartilage (Swahn et al., 2022) used as a reference for BayesPrism deconvolution.

**B,** Feature plots of representative marker genes used for cell type identification in the reference dataset.

**Supplementary Fig. S8 | Construction of the single-cell RNA-seq reference of meniscus for deconvolution.**

**A,** UMAP visualization of the re-analyzed public single-cell RNA-seq dataset of meniscus (Swahn et al., 2022) used as a reference for BayesPrism deconvolution.

**B,** Feature plots of representative marker genes used for cell type identification in the reference dataset.

**Supplementary Fig. S9 | Epigenetic binding and expression profile of ETS1.**

**A**, Visualization of ETS1 ChIP-seq peaks at the *EXOSC6* genomic locus, retrieved from the ChIP-Atlas database.

**B**, Single-nucleus gene expression levels of *ETS1* within the meniscus, derived from snMultiome sequencing data.

## Supplementary Tables

**Supplementary Table S1. | Genome and RNA Datasets, Sex, Race, and Disease Severity.**

**Supplementary Table S2. | Differentially expressed genes for cartilage.**

**Supplementary Table S3. | Differentially expressed genes for meniscus.**

**Supplementary Table S4. | GWAS colocalized genes with RTC > 0.9.**

**Supplementary Table S5. | GWAS colocalized genes with Coloc PP.H4 > 0.75.**

**Supplementary Table S6. | Sexual differences of eQTL signals.**

**Supplementary Table S7. | Differences of eQTL signals by disease severity.**

**Supplementary Table S8. | Promoter AI and expression changes of cartilage and meniscus.**

**Supplementary Table S9. | Prediction of cell-type-specific enhancers via Single-nuclei Multiome analysis.**

**Supplementary Table S10. | GWAS colocalized genes for each cell cluster (RTC > 0.9).**

**Supplementary Table S11. | GWAS colocalized genes for each cell cluster (Coloc PPH4 > 0.75).**

**Supplementary Table S12. | Finemapped eQTL SNVs in promoter regions.**

**Supplementary Table S13. | Finemapped eQTL SNVs in enhancer regions.**

**Supplementary Table S14. | Candidate therapeutic compounds for the reversal of disease-associated eQTLs and transcriptomic signatures.**

