## Supplementary Figures for "Integrated mapping of human meniscus and cartilage eQTLs reveals shared and distinct osteoarthritis genetic drivers"

Supplementary Fig. S1

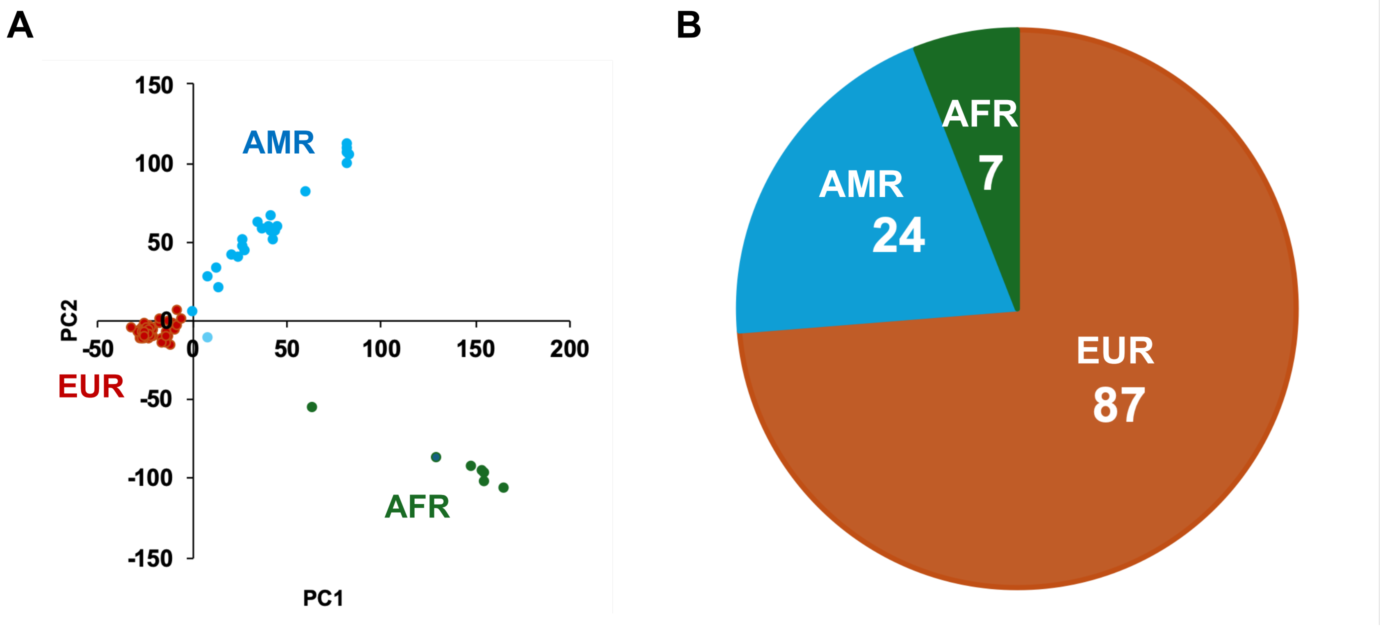

### Supplementary Fig. S2

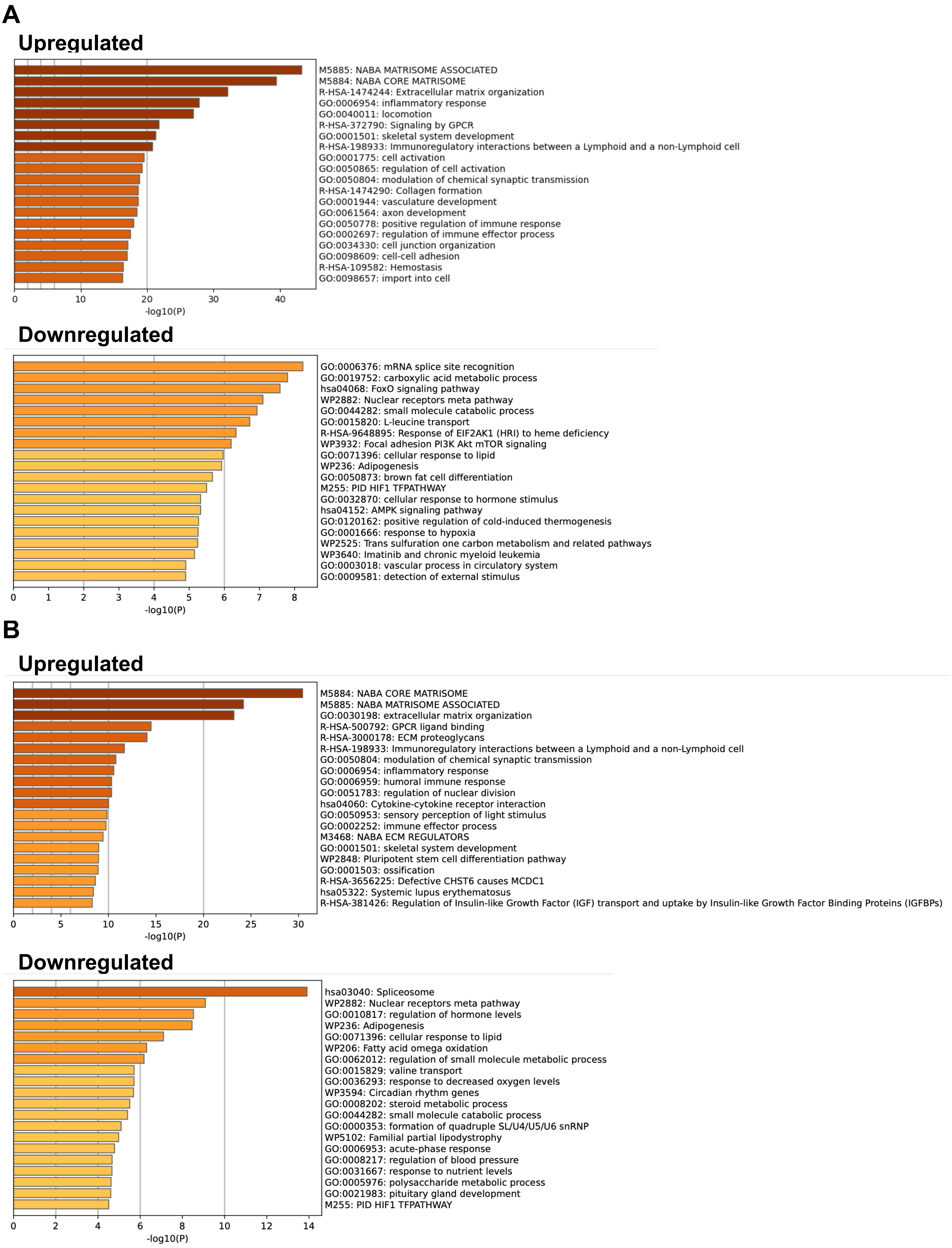

### Supplementary Fig. S3

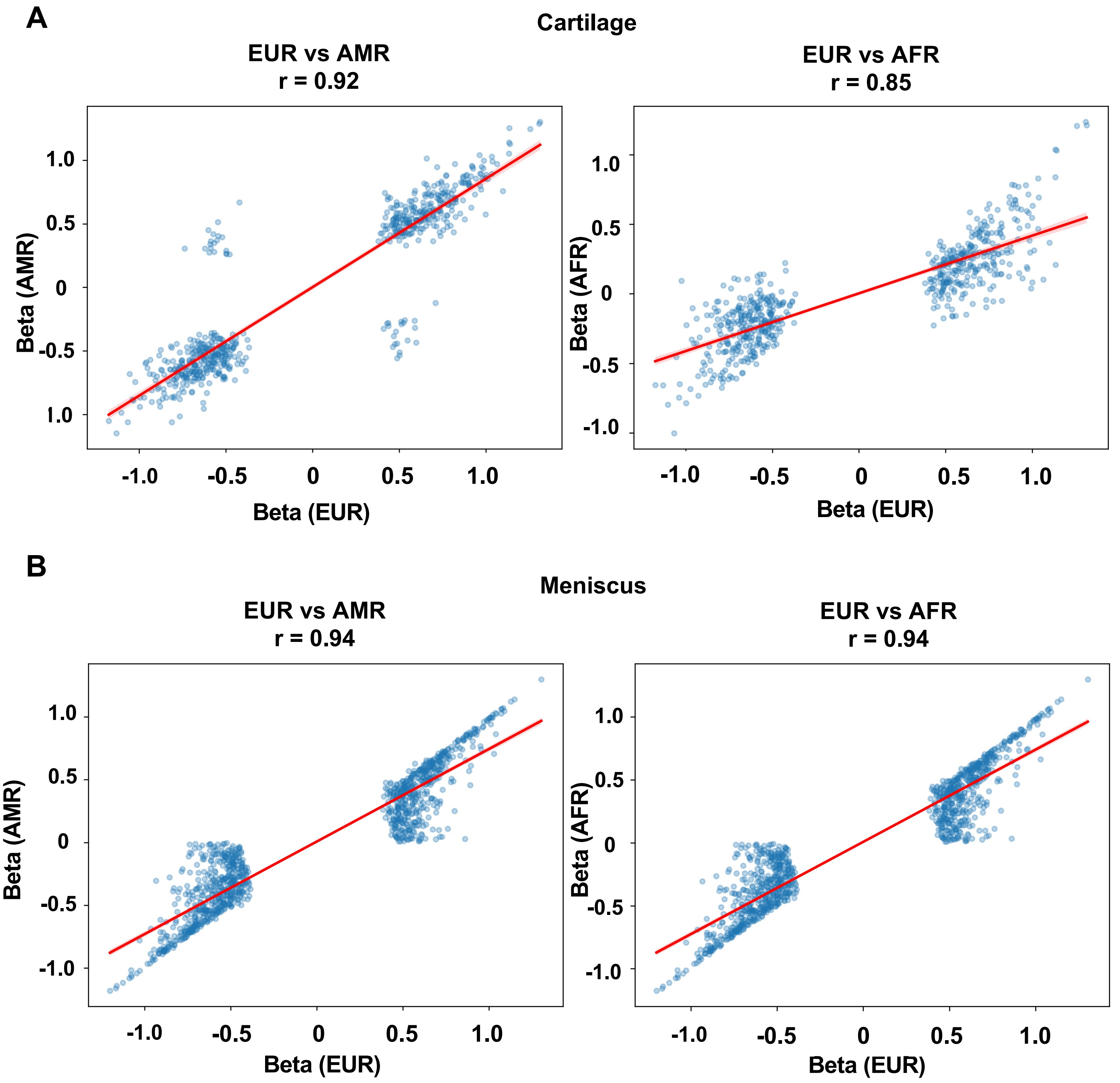

#### Supplementary Fig. S4

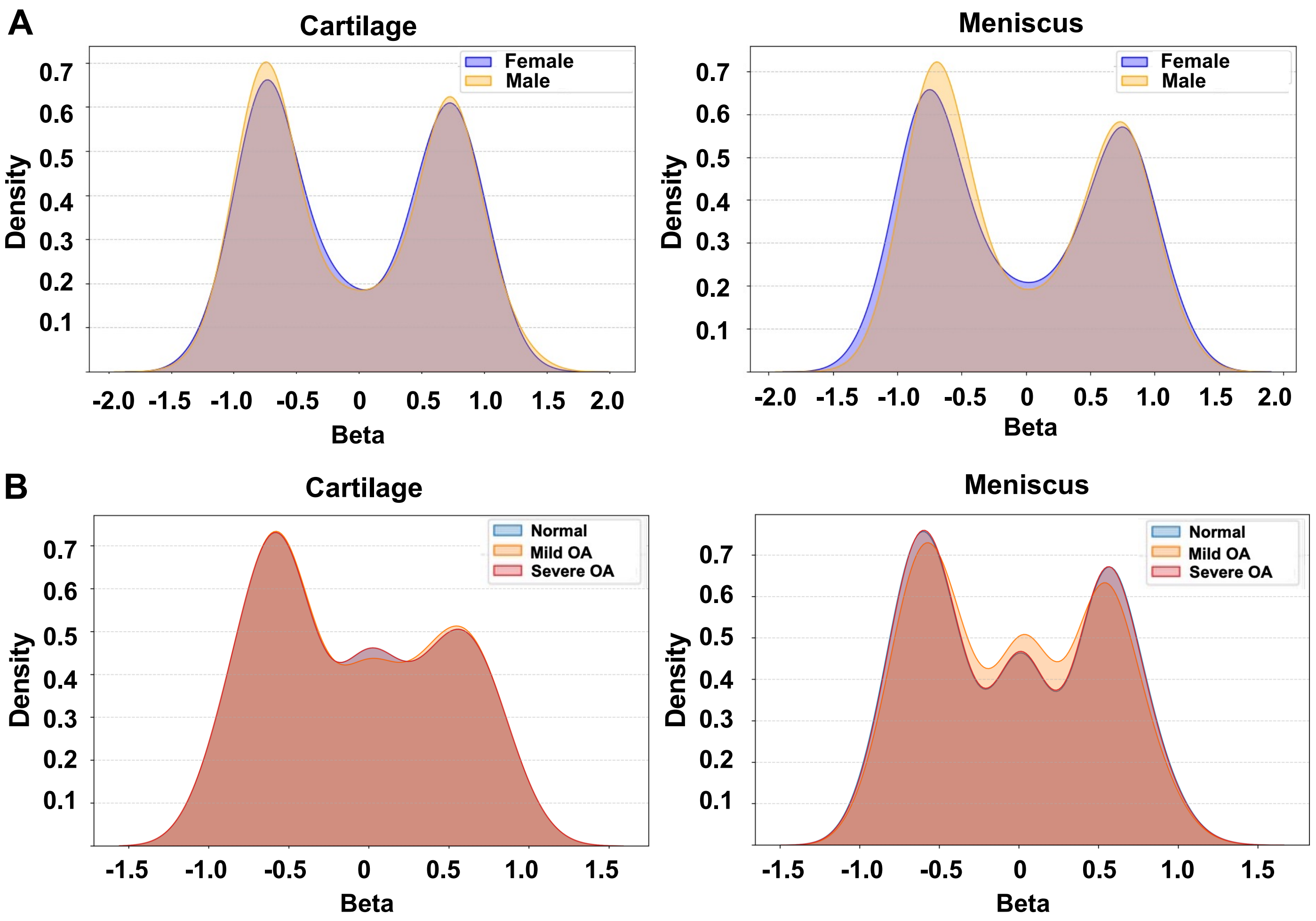

#### Supplementary Fig. S5

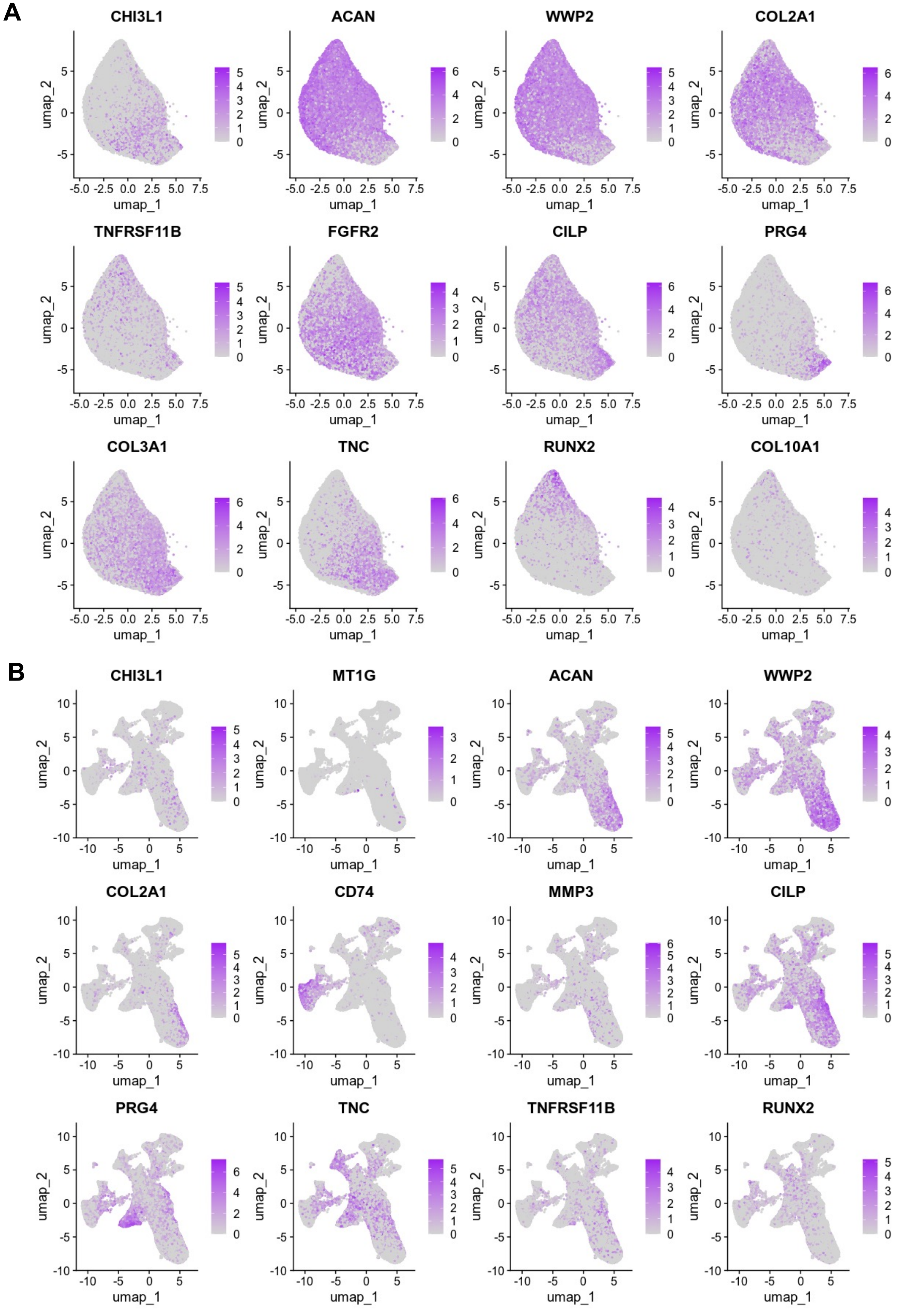

#### Supplementary Fig. S6

**A**

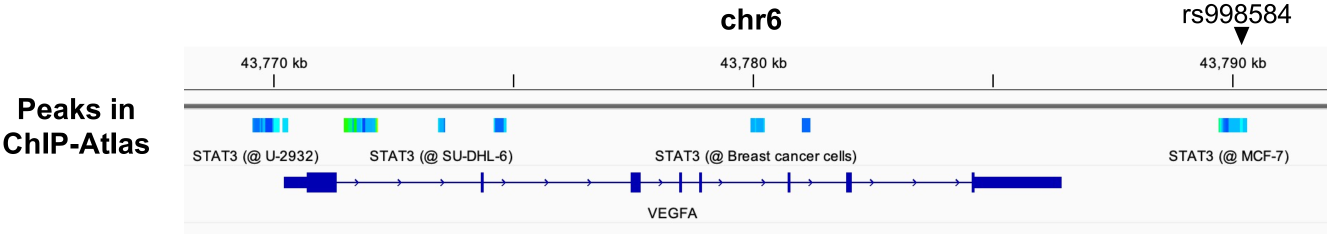


**B**

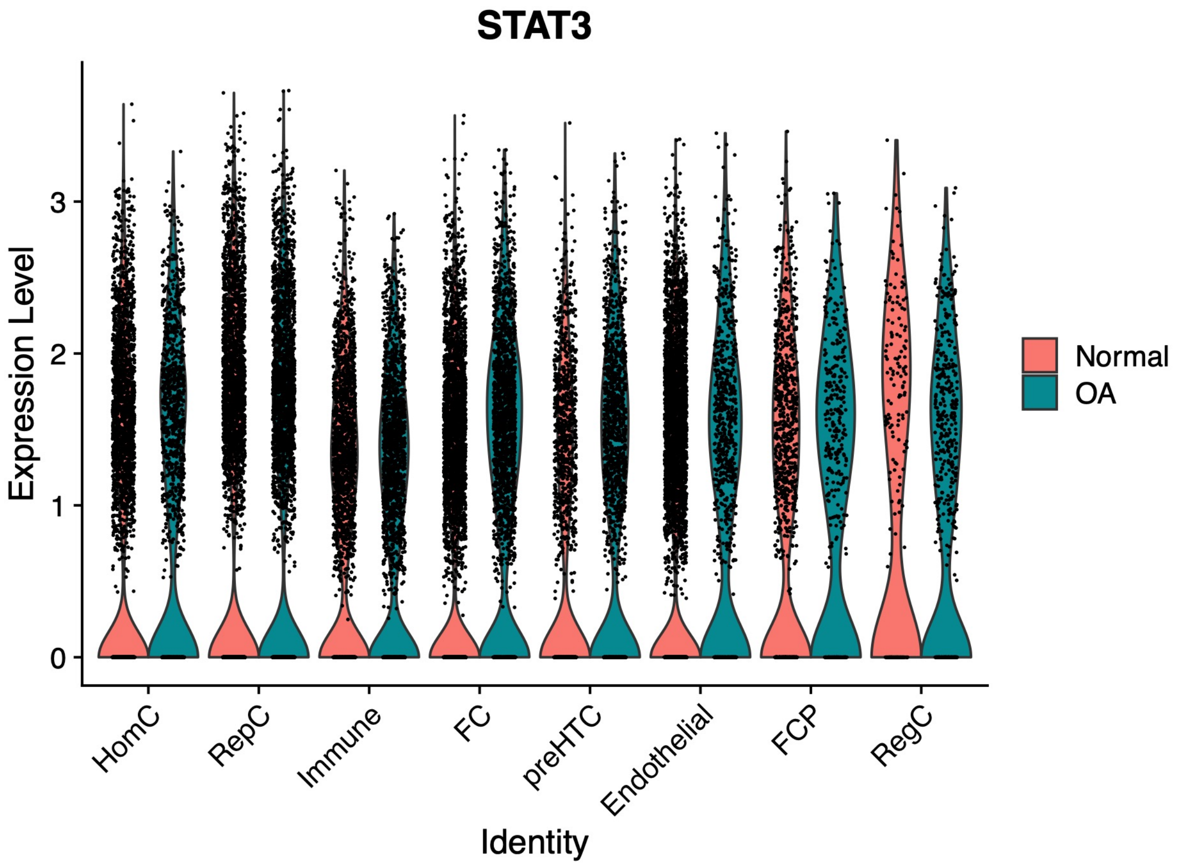

### Supplementary Fig. S7

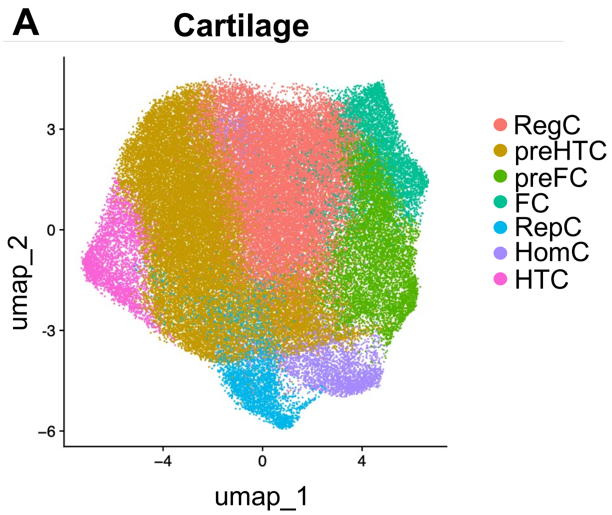


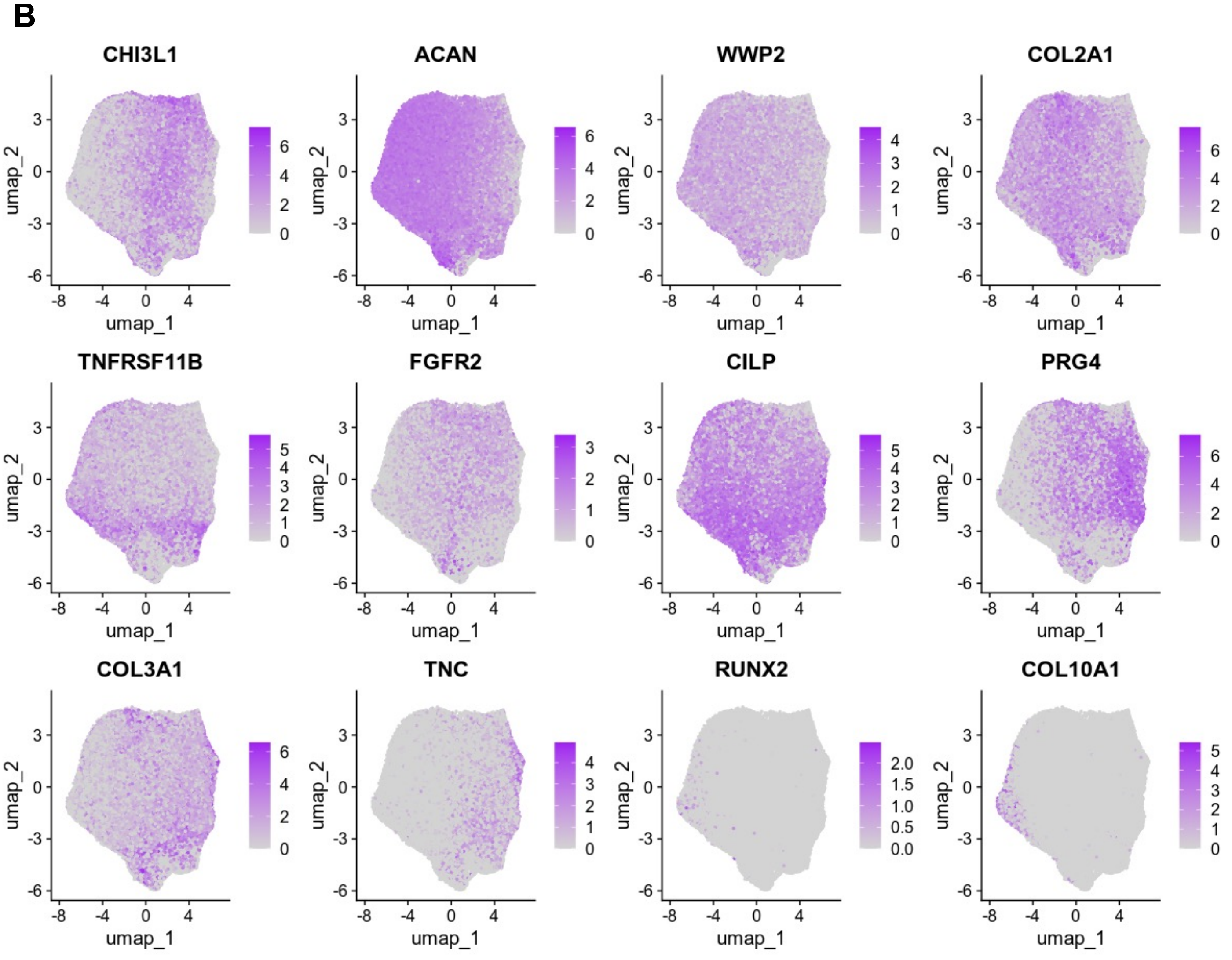

#### Supplementary Fig. S8

A

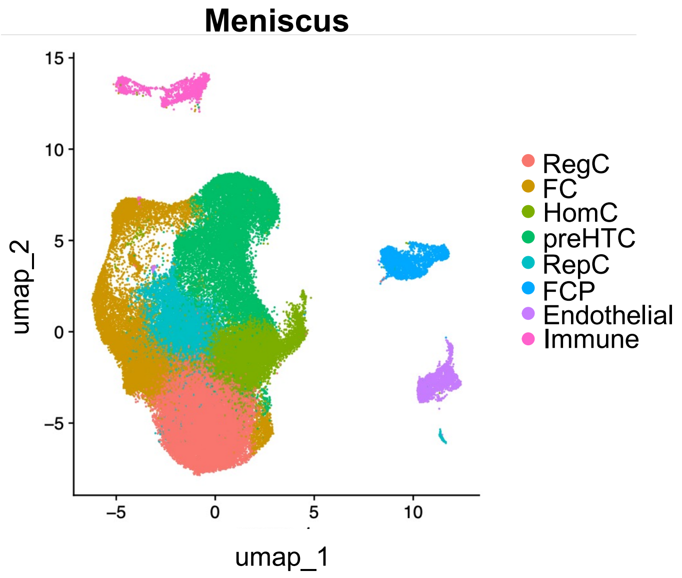


B

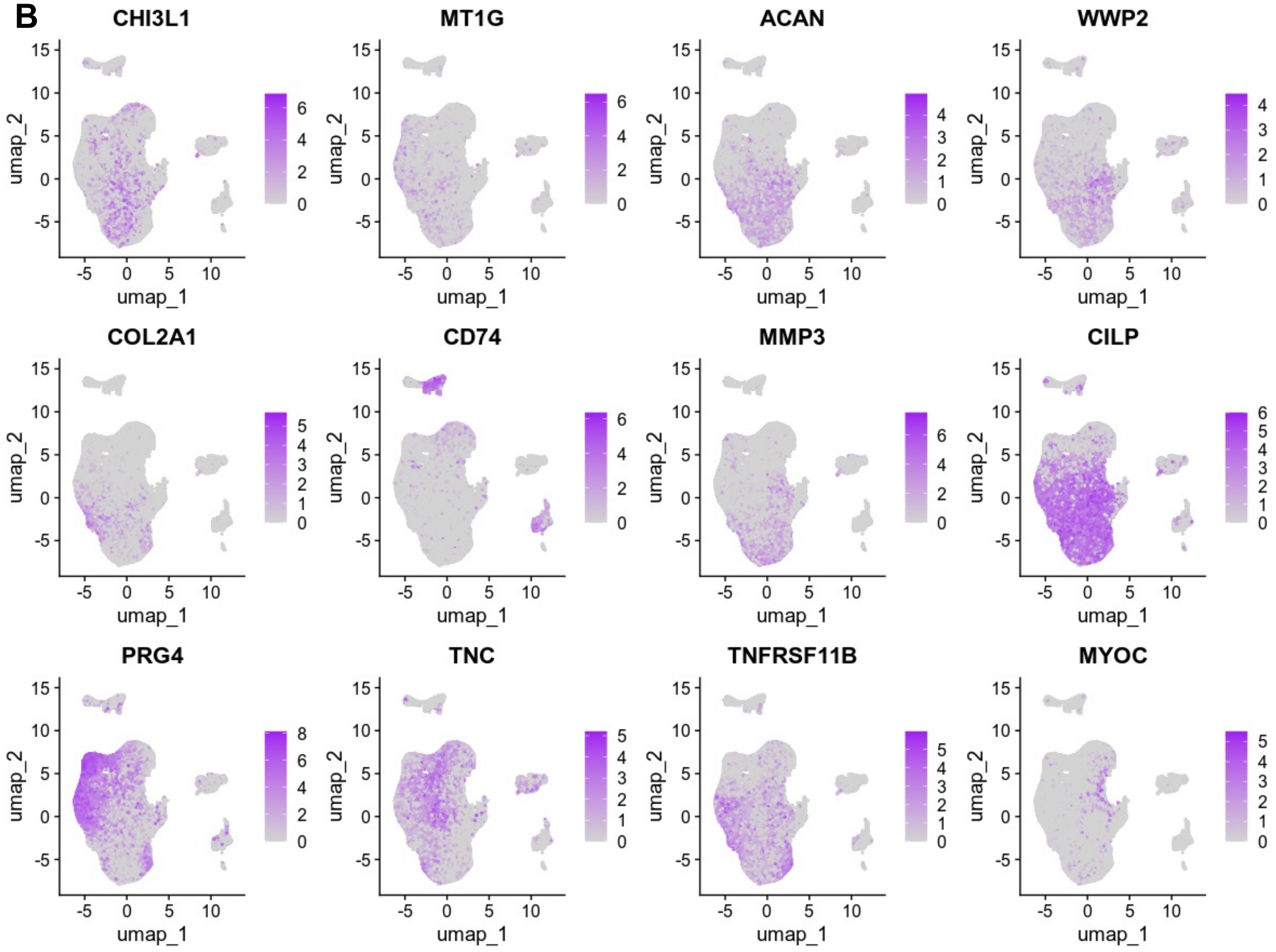

#### Supplementary Fig. S9

**A**

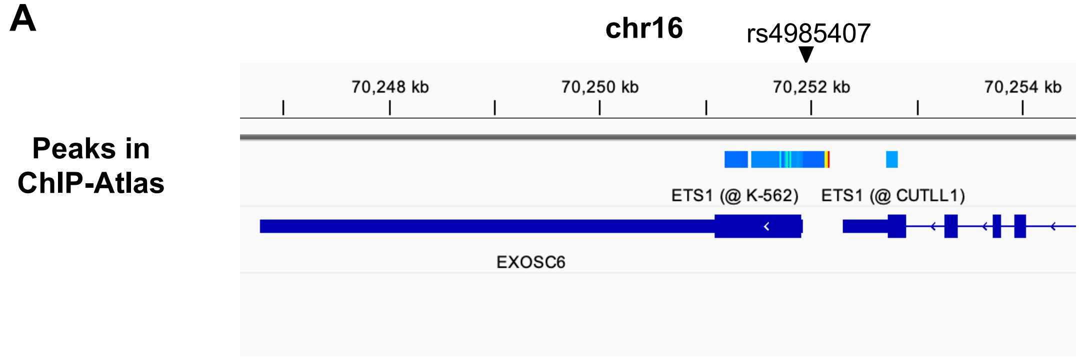


**B**

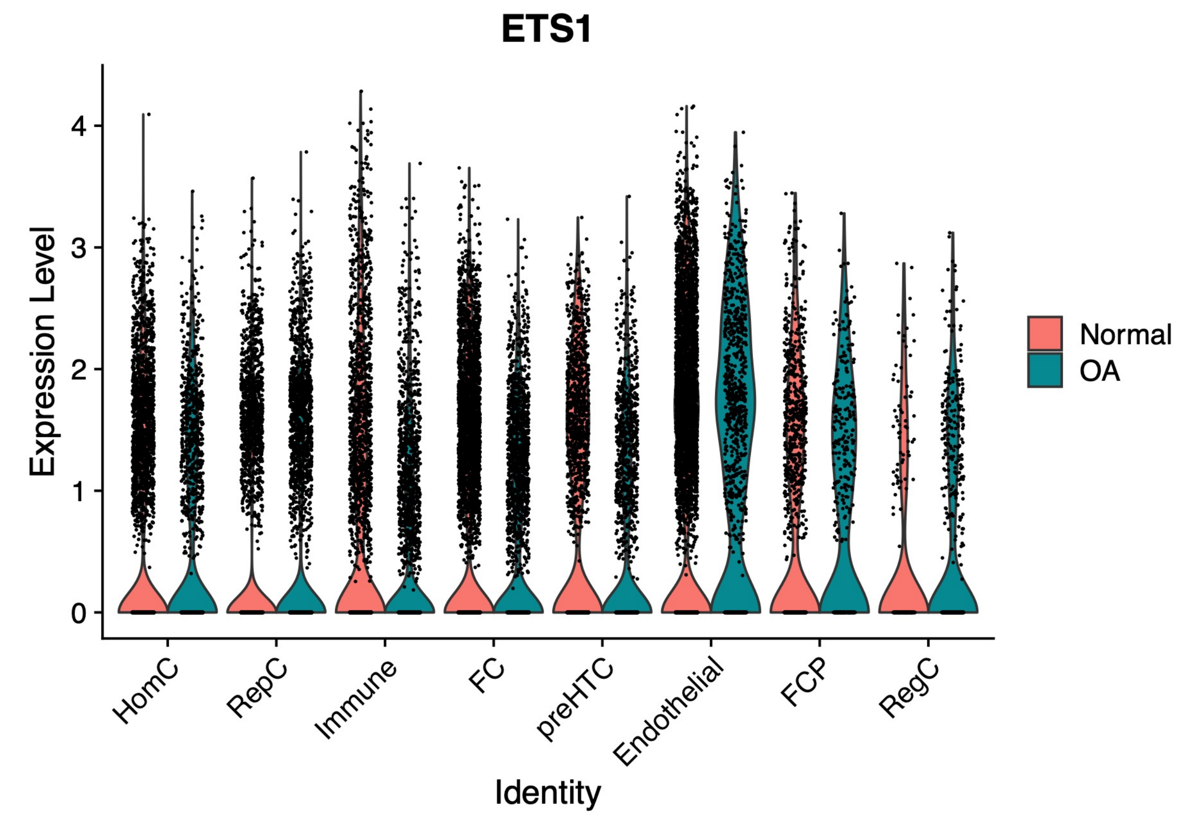
